# Interleukin-6 Receptor Antagonists in Critically Ill Patients with Covid-19 – Preliminary report

**DOI:** 10.1101/2021.01.07.21249390

**Authors:** The REMAP-CAP Investigators, Anthony C. Gordon, Paul R. Mouncey, Farah Al-Beidh, Kathryn M. Rowan, Alistair D. Nichol, Yaseen M. Arabi, Djillali Annane, Abi Beane, Wilma van Bentum-Puijk, Lindsay R. Berry, Zahra Bhimani, Marc J.M. Bonten, Charlotte A. Bradbury, Frank M. Brunkhorst, Adrian Buzgau, Allen C. Cheng, Michelle A. Detry, Eamon J. Duffy, Lise J. Estcourt, Mark Fitzgerald, Herman Goossens, Rashan Haniffa, Alisa M. Higgins, Thomas E. Hills, Christopher M. Horvat, Francois Lamontagne, Patrick R. Lawler, Helen L. Leavis, Kelsey M. Linstrum, Edward Litton, Elizabeth Lorenzi, John C. Marshall, Florian B. Mayr, Danny McAuley, Anna McGlothlin, Shay P McGuinness, Bryan J. McVerry, Stephanie K. Montgomery, Susan C. Morpeth, Srinivas Murthy, Katrina Orr, Rachael L. Parke, Jane C. Parker, Asad E. Patanwala, Ville Pettilä, Emma Rademaker, Marlene S. Santos, Christina T. Saunders, Christopher W. Seymour, Manu Shankar-Hari, Wendy I. Sligl, Alexis F. Turgeon, Anne M. Turner, Frank L. van de Veerdonk, Ryan Zarychanski, Cameron Green, Roger J. Lewis, Derek C. Angus, Colin J. McArthur, Scott Berry, Steve A. Webb, Lennie P.G. Derde

**Affiliations:** Imperial College London, London, United Kingdom; Imperial College Healthcare NHS Trust, St. Mary’s Hospital, London, United Kingdom; Intensive Care National Audit & Research Centre (ICNARC), London, United Kingdom; Monash University, Melbourne, Australia; University College Dublin, Dublin, Ireland; Alfred Health, Melbourne, Australia; King Saud bin Abdulaziz University for Health Sciences and King Abdullah International Medical Research Center, Riyadh, Kingdom of Saudi Arabia; Hospital Raymond Poincaré (Assistance Publique Hôpitaux de Paris), Garches, France; Université Versailles SQY - Université Paris Saclay, Montigny-le-Bretonneux, France; Université Paris Saclay - UVSQ – INSERM, Garches, France; University of Oxford, Oxford, United Kingdom; University Medical Center Utrecht, Utrecht, The Netherlands; Berry Consultants, Austin, United States; St. Michael’s Hospital Unity Health, Toronto, Canada; University of Bristol, Bristol, United Kingdom; Jena University Hospital, Jena, Germany; Auckland District Health Board, Auckland, New Zealand; NHS Blood and Transplant, Oxford, United Kingdom; University of Antwerp, Wilrijk, Belgium; University of Oxford, Bangkok, Thailand; University College London Hospital, London, United Kingdom; National Intensive Care Surveillance (NICST), Colombo, Sri Lanka; Auckland City Hospital, Auckland, New Zealand; Medical Research Institute of New Zealand (MRINZ), Wellington, New Zealand; UPMC Children’s Hospital of Pittsburgh, Pittsburgh, United States; Université de Sherbrooke, Sherbrooke, Quebec, Canada; University Health Network, Toronto, Canada; University of Toronto, Toronto, Canada; University of Pittsburgh, Pittsburgh, United States; Fiona Stanley Hospital, Perth, Australia; University of Western Australia, Perth, Australia; Queen’s University Belfast, Belfast, Northern Ireland; Royal Victoria Hospital, Belfast, Northern Ireland; Middlemore Hospital, Auckland, New Zealand; University of British Columbia, Vancouver, Canada; University of Auckland, Auckland, New Zealand; University of Sydney, Sydney, Australia; Royal Prince Alfred Hospital, Sydney, Australia; University of Helsinki and Helsinki University Hospital, Helsinki, Finland; King’s College London, London, United Kingdom; Guy’s and St Thomas’ NHS Foundation Trust, London, United Kingdom; University of Alberta, Edmonton, Canada; Université Laval, Québec City, Canada; Radboudumc, Nijmegen, The Netherlands; University of Manitoba, Winnipeg, Canada; Harbor-UCLA Medical Center, Torrance, CA, United States; St John of God Hospital, Subiaco, Australia

**Keywords:** Adaptive platform trial, intensive care, pneumonia, pandemic, Covid-19, tocilizumab, sarilumab, interleukin-6

## Abstract

**Background:** The efficacy of interleukin-6 receptor antagonists in critically ill patients with coronavirus disease 2019 (Covid-19) is unclear.

**Methods:** We evaluated tocilizumab and sarilumab in an ongoing international, multifactorial, adaptive platform trial. Adult patients with Covid-19, within 24 hours of commencing organ support in an intensive care unit, were randomized to receive either tocilizumab (8mg/kg) or sarilumab (400mg) or standard care (control). The primary outcome was an ordinal scale combining in-hospital mortality (assigned −1) and days free of organ support to day 21. The trial uses a Bayesian statistical model with pre-defined triggers to declare superiority, efficacy, equivalence or futility.

**Results:** Tocilizumab and sarilumab both met the pre-defined triggers for efficacy. At the time of full analysis 353 patients had been assigned to tocilizumab, 48 to sarilumab and 402 to control. Median organ support-free days were 10 (interquartile range [IQR] −1, 16), 11 (IQR 0, 16) and 0 (IQR −1, 15) for tocilizumab, sarilumab and control, respectively. Relative to control, median adjusted odds ratios were 1.64 (95% credible intervals [CrI] 1.25, 2.14) for tocilizumab and 1.76 (95%CrI 1.17, 2.91) for sarilumab, yielding >99.9% and 99.5% posterior probabilities of superiority compared with control. Hospital mortality was 28.0% (98/350) for tocilizumab, 22.2% (10/45) for sarilumab and 35.8% (142/397) for control. All secondary outcomes and analyses supported efficacy of these IL-6 receptor antagonists.

**Conclusions:** In critically ill patients with Covid-19 receiving organ support in intensive care, treatment with the IL-6 receptor antagonists, tocilizumab and sarilumab, improved outcome, including survival. (ClinicalTrials.gov number: NCT02735707)

## Background

Globally, there have been over 79 million reported cases of Coronavirus Infectious Disease 2019 (Covid-19) with over 1.75 million deaths.^1^ Only corticosteroids are known to improve survival for severely ill patients.^2^ The benefit from corticosteroids in critically ill patients supports the concept that an excessive host inflammatory response is responsible for much of the morbidity and mortality from Covid-19.

Interleukin-6 (IL-6) is released in response to infection and stimulates inflammatory pathways as part of the acute phase response. Tocilizumab and sarilumab are monoclonal antibodies that inhibit both membrane-bound and soluble IL-6 receptors and are used to treat inflammatory conditions, such as rheumatoid arthritis, and cytokine release syndrome after chimeric antigen receptor T-cell (CAR-T) therapy (tocilizumab). Their clinical use has been described in Covid-19,^3-5^ however, randomized controlled trials to date have been inconclusive.^6-10^

We investigated the effectiveness of tocilizumab and sarilumab on survival and organ support in critically ill patients with Covid-19 in the Randomized, Embedded, Multifactorial Adaptive Platform Trial for Community-Acquired Pneumonia (REMAP-CAP).

## Methods

### Trial Design and Oversight

REMAP-CAP is an international, adaptive platform trial designed to determine best treatment strategies for patients with severe pneumonia in both pandemic and non-pandemic settings. REMAP-CAP’s design and first results, regarding corticosteroids in Covid-19, were published previously.^11,12^

Patients eligible for the platform are assessed for eligibility and potentially randomized to multiple interventions across multiple domains. A ‘domain’ covers a common therapeutic area (e.g., antiviral therapy) and contains two or more interventions (including control e.g. ‘no antiviral’). Patients are randomized to one intervention in each domain for which they are eligible. The REMAP-CAP trial is defined by a master (‘core’) protocol with individual appendices for each domain, regional governance and adaptations for a declared pandemic. The trial is overseen by a blinded International Trial Steering Committee (ITSC) and an unblinded independent Data and Safety Monitoring Board (DSMB). The trial is approved by relevant regional ethics committees (see Supplementary Appendix for more detail) and is conducted in accordance with Good Clinical Practice guidelines and the principles of the Declaration of Helsinki. Written or verbal informed consent, in accordance with regional legislation, is obtained from all patients or their surrogates.

The trial has multiple funders, internationally, with multiple regional sponsors. Roche Products Ltd and Sanofi supported the trial through provision of tocilizumab and sarilumab in the United Kingdom. The funders, sponsors, and Roche and Sanofi had no role in designing the trial, analyzing data, writing the manuscript, or making the decision to submit for publication. All authors vouch for the data and analyses, as well as for the fidelity of this report to the trial protocol and statistical analysis plan.

### Participants

Critically ill patients, aged ≥18 years, with suspected or confirmed Covid-19, admitted to an intensive care unit (ICU) and receiving respiratory or cardiovascular organ support were classified as severe state and were eligible for enrollment in the Covid-19 Immune Modulation Therapy domain. Exclusion criteria included presumption that death was imminent with lack of commitment to full support, and prior participation in REMAP-CAP within 90 days. Additional exclusion criteria, specific for the Immune Modulation Therapy domain, are listed in the Supplementary Appendix.

### Randomization

The Immune Modulation Therapy domain included five interventions: two IL-6 receptor antagonists, tocilizumab and sarilumab; an IL-1 receptor antagonist, anakinra; and interferon beta-1a; as well as control (no immune modulation). Investigators at each site selected *a priori* at least two interventions, one of which had to be control, to which patients would be randomized. Participants were randomized via centralized computer program to each intervention (available at the site) starting with balanced assignment for tocilizumab, sarilumab or control (e.g. 1:1 if two interventions available, 1:1:1 if three interventions available).

Tocilizumab, at a dose of 8mg/kg of actual body weight (up to a maximum of 800mg), was administered as an intravenous infusion over one hour; this dose could be repeated 12-24 hours later at the discretion of the treating clinician. Sarilumab, 400mg, was administered as an intravenous infusion once only. All investigational drugs were dispensed by local pharmacies and were open-label.

### Procedures

Other aspects of patient management were provided per each site’s standard of care. In addition to assignments in this domain, participants could be randomized to other interventions within other domains, depending on domains active at the site, patient eligibility, and consent (see www.remapcap.org). Randomization to the Corticosteroid domain for Covid-19 closed on June 17, 2020.^12^ Thereafter, corticosteroids were allowed as per recommended standard of care.

Although clinical staff were aware of individual patient intervention assignment, neither they nor the ITSC were provided any information about aggregate patient outcomes.

### Outcome Measures

The primary outcome was respiratory and cardiovascular organ support-free days up to day 21. In this composite ordinal outcome, all deaths within hospital are assigned the worst outcome (–1). Among survivors, respiratory and cardiovascular organ support-free days are calculated up to day 21, such that a higher number represents faster recovery. This outcome was used in a recent Food and Drug Administration approved trial and 1.5 days was considered a minimally clinically important difference.^13^ Secondary outcomes were all pre-specified and details are in the Supplementary Appendix.

### Statistical Analysis

REMAP-CAP uses a Bayesian design with no maximum sample size. Regular, interim analyses are conducted and randomization continues, potentially with response-adaptive randomization with preferential assignment to those interventions that appear most favorable, until a pre-defined statistical trigger is met.

The primary analysis was generated from a Bayesian cumulative logistic model, which calculated posterior probability distributions of the 21-day organ support-free days (primary outcome) based on evidence accumulated in the trial and assumed prior knowledge in the form of a prior distribution. Prior distributions for individual treatment effects were neutral. The primary model adjusted for location (site, nested within country), age (categorized into six groups), sex, and time-period (two-week epochs). The model contained treatment effects for each intervention within each domain and pre-specified treatment-by-treatment interactions across domains. The treatment effects for tocilizumab and sarilumab were “nested” in the model with a hierarchical prior distribution sharing a common mean and variance. This prior structure facilitates dynamic borrowing between the two IL-6 receptor antagonists that borrows more information when the observed effects are similar and less when they are different.^14^

The primary analysis was conducted on all severe state patients with Covid-19 randomized to any domain up to November 19, 2020 (and with complete follow-up). The inclusion of additional patients enrolled outside the Immune Modulation Therapy domain allows maximal incorporation of all information, providing the most robust estimation of the coefficients of all covariates, as per the principle of the REMAP-CAP design.^11,12^ Importantly, not all patients were eligible for all domains nor for all interventions (dependent on active domains and interventions at the site, eligibility criteria, and patient/surrogate consent). Therefore, the model included covariate terms reflecting each patient’s domain eligibility, such that the estimate of an intervention’s effectiveness, relative to any other intervention within that domain, was generated from those patients that might have been eligible to be randomized to those interventions within the domain.

The cumulative log odds for the primary outcome was modeled such that a parameter >0 reflects an increase in the cumulative log odds for the organ support-free days outcome, implying benefit. There was no imputation of missing outcomes. The model was fit using a Markov Chain Monte Carlo algorithm that drew iteratively (10,000 draws) from the joint posterior distribution, allowing calculation of odds ratios with their 95% credible intervals (CrI) and the probability that each intervention (including control) was optimal in the domain, that an intervention was superior compared with control (efficacy), that two non-control interventions were equivalent, or an intervention was futile compared with control. An odds ratio >1 represents improved survival and/or more organ support-free days. The pre-defined statistical triggers for trial conclusions and disclosure of results were: a >99% posterior probability that an intervention was optimal compared with all other interventions; an inferiority conclusion if <0.25% posterior probability that an intervention was optimal; an intervention efficacy if >99% posterior probability the odds ratio was >1 compared with control; intervention futility if <5% posterior probability the odds ratio was >1.2 compared with control, or equivalence if >90% probability the odds ratio was between 1/1.2 and 1.2 for two non-control interventions.

Analysis of the primary outcome was then repeated in a second model using only data from those patients enrolled in domains that had stopped and were unblinded at the time of analysis with no adjustment for assignment in other ongoing domains. The secondary outcomes were also analyzed in this second model. One subgroup analysis, based on terciles of serum C-reactive protein (CRP) at inclusion, was pre-specified. Further details of all analyses are provided in the Supplementary Appendix. Pre-specified analyses are listed in the statistical analysis plan. Data management and summaries were created using R version 3.6.0, the primary analysis was computed in R version 4.0.0 using the rstan package version 2.21.1. Additional data management and analyses were performed in SQL 2016, SPSS version 26, and Stata version 14.2.

## Results

These are preliminary results. As further follow-up and analysis continues, minor changes may occur.

The first patient with Covid-19 was enrolled into REMAP-CAP on March 9, 2020 and the first patient randomized in the Immune Modulation Therapy domain on April 19 as tocilizumab became available. Sarilumab only became available later. At a scheduled interim analysis, the independent DSMB reported that tocilizumab had met the statistical trigger for efficacy (posterior probability 99.75%, odds ratio 1.87, 95%CrI 1.20, 2.76) based on an interim analysis of patients as of October 28. As per protocol, further assignment to control closed on November 19 with randomization continuing between different active immune modulation interventions. At this time, 2,046 patients had been randomized in at least one domain in the severe disease state of REMAP-CAP and 895 had been randomized in the Immune Modulation Therapy domain (366 to tocilizumab, 48 to sarilumab, 412 to control and 69 to other interventions within the domain) in 113 sites across six countries (Figure 1). Thirty patients subsequently withdrew consent, and 11 patients had missing primary outcome data. Following a subsequent interim analysis, the DSMB reported that sarilumab had also met the statistical trigger for efficacy and so these results are also reported.

**Figure 1.**
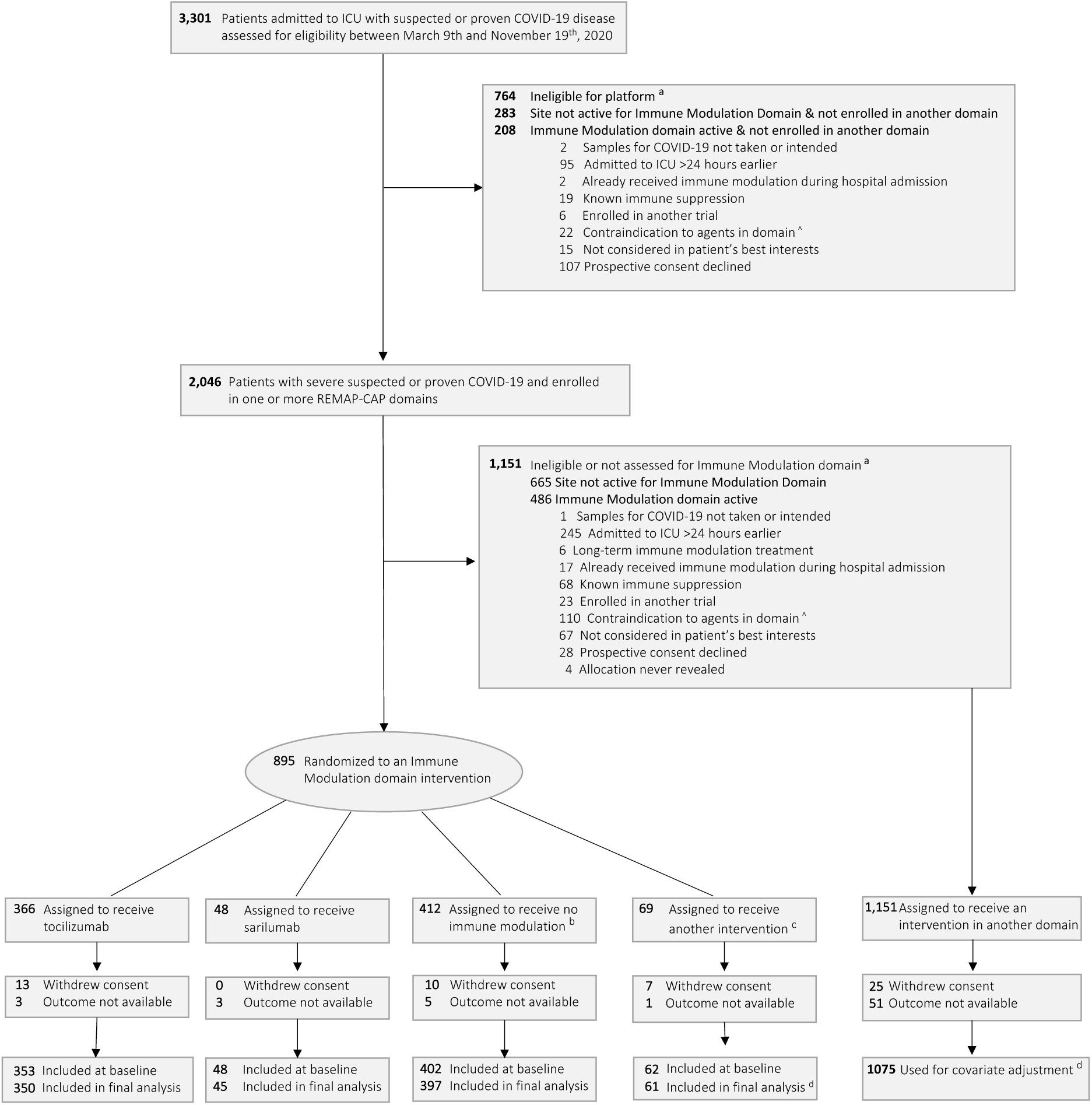
Screening, enrollment, randomization and inclusion in analysis. a = Patients could meet more than one ineligibility criterion. Full details are provided in the supplement b = only includes patients when tocilizumab and /or sarilumab was a randomization option c = other interventions includes anakinra, interferon β1a, and no immune modulation when tocilizumab and / or sarilumab was not available as a randomization option d = The primary analysis of alternative interventions within the immune modulation domain is estimated from a model that adjusts for patient factors and for assignment to interventions in other domains. To obtain the most reliable estimation of the effect of these patient factors and of other interventions on the primary outcome, all patients enrolled in the severe COVID-19 cohort (for whom there is consent and follow-up) are included. Importantly, however, the model also factors eligibility for the immune modulation domain and its interventions, such that the final estimate of an immune modulation domain intervention’s effectiveness relative to any other within that domain is generated from those patients that might have been eligible to be randomized to those interventions within the domain. ^ Contraindications include hypersensitivity, raised ALT/AST, or thrombocytopenia, or pregnancy

### Patients

Baseline characteristics were balanced across intervention groups and typical of a critically ill population with Covid-19 (Table 1). All but three patients were receiving respiratory support at the time of randomization, including high flow nasal oxygen (28.8%), non-invasive (41.5%) and invasive (29.4%) mechanical ventilation. The majority of patients (n=707) were enrolled after June 17 and the announcement of the dexamethasone result from the RECOVERY trial^15^ and of these patients, 93.3% (610/654) were treated with corticosteroids at enrollment or within the following 48 hours. Of the 158 patients recruited before June 17, 107 were randomized in the previously published Corticosteroid domain within REMAP-CAP, 41 allocated to a seven-day course of hydrocortisone and 39 to shock-dependent hydrocortisone.^12^ Remdesivir use was recorded in 32.8% (265/807) of patients.

**Table 1.** Baseline Characteristics of Participants in the Immune Modulation Therapy domain*.

| Characteristic | Tocilizumab<br>(N=353) | Sarilumab<br>(N=48) | Control <sup>a</sup><br>(N=402) | All participants in<br>the Immune<br>Modulation<br>Therapy domain <sup>b</sup><br>(N=865) |
| --- | --- | --- | --- | --- |
| Age - mean (SD), years | 61.5 (12.5) | 63.4 (13.4) | 61.1 (12.8) | 61.4 (12.7) |
| Male Sex - n (%) | 261 (73.9) | 39 (81.3) | 283 (70.4) | 629 (72.7) |
| Race/Ethnicity <sup>c</sup> -<br>White - n/N (%) | 160/228 (70.2) | 29/39 (74.4) | 206/279 (73.8) | 420/580 (72.4) |
| Asian - n/N (%) | 41/228 (18.0) | 8/39 (20.5) | 47/279 (16.9) | 99/580 (17.1) |
| Black - n/N (%) | 12/228 (5.3) | 1/39 (2.6) | 9/279 (3.2) | 23/580 (4.0) |
| Mixed - n/N (%) | 2/228 (0.9) | 0/39 (0.0) | 5/279 (1.8) | 7/580 (1.2) |
| Other - n/N (%) | 13/228 (5.7) | 1/39 (2.6) | 12/279 (4.3) | 31/580 (5.3) |
| Body mass index <sup>d</sup> - median<br>(IQR), kg/m<sup>2</sup> | 30.5 (26.9-34.9)<br>(n=342) | 29.2 (26.0-33.8)<br>(n=39) | 30.9 (27.1-34.9)<br>(n=377) | 30.5 (26.8-34.9)<br>(n=815) |
| APACHE II score <sup>e</sup> - median (IQR) | 13 (8-19) (n=337) | 10 (7-16) (n=42) | 12 (8-18) (n=381) | 12.5 (8-19) (n=820) |
| Confirmed SARS-CoV2 infection<sup>f</sup><br>– n/N (%) | 284/345 (82.3) | 44/47 (93.6) | 334/394 (84.8) | 715/847 (84.4) |
| Pre-existing conditions – n/N<br>(%) |  |  |  |  |
| Diabetes mellitus | 123/349 (35.2) | 13/48 (27.1) | 150/401 (37.4) | 304/860 (35.4) |
| Kidney disease | 30/312 (9.6) | 4/45(8.9) | 43/372 (11.6) | 81/789 (10.3) |
| Respiratory disease<sup>g</sup> | 82/349 (23.5) | 15/48 (31.3) | 98/401 (24.4) | 206/860 (24.0) |
| Immunosuppressive disease | 8/348 (2.3) | 0/48 (0.0) | 14/401 (3.5) | 25/859 (2.9) |
| Chronic immunosuppressive<br>therapy | 3/349 (0.9) | 1/48 (2.1) | 6/401 (1.5) | 12/860 (1.4) |
| Severe cardiovascular disease | 34/339 (10.0) | 1/48 (2.1) | 47/395 (11.9) | 86/844 (10.2) |
| Liver cirrhosis/failure | 2/339 (0.6) | 0/48 (0.0) | 1/395 (0.3) | 5/844 (0.6) |
| Time to enrollment - median<br>(IQR) |  |  |  |  |
| From hospital admission -<br>days | 1.2 (0.8-2.8) | 1.4 (0.9-2.8) | 1.2 (0.8-2.8) | 1.2 (0.8-2.8) |
| From ICU admission - hours | 13.1 (6.6-19.0) | 16.0 (11.4-20.8) | 14.0 (6.8-19.5) | 13.6 (6.6-19.4) |
| Acute respiratory support |  |  |  |  |
| None/supplemental oxygen<br>only | 1/353 (0.3) | 0/48 (0.0) | 2/402 (0.5) | 3/865 (0.4) |
| High flow nasal cannula | 101/353 (28.6) | 17/48 (35.4) | 110/402 (27.4) | 249/865 (28.8) |
| Non-invasive ventilation only | 147/353 (41.6) | 23/48 (47.9) | 169/402 (42.0) | 359/865 (41.5) |
| Invasive mechanical ventilation | 104/353 (29.5) | 8/48 (16.7) | 121/402 (30.1) | 254/865 (29.4) |
| Vasopressor support | 63/353 (17.9) | 4/48 (8.3) | 79/402 (19.7) | 163/865 (18.8) |
| PaO<sub>2</sub>/FiO<sub>2</sub> – median (IQR) | 115 (89-162)<br>(n=335) | 126 (99-157) | 118 (89-169)<br>(n=354) | 116.5 (89-165)<br>(n=780) |
| Median laboratory values (IQR)<sup>g</sup> |  |  |  |  |
| C-reactive protein, µg/mL | 150 (85-221)<br>(n=207) | 136 (105-204)<br>(n=37) | 130 (71-208)<br>(n=244) | 136 (79-208)<br>(n=533) |
| D-dimer, ng/mL | 832 (461-1763)<br>(n=159) | 828 (355-1435)<br>(n=20) | 1010 (500-2115)<br>(n=172) | 910 (480-1916)<br>(n=385) |
| Ferritin, ng/mL | 912 (525-1590)<br>(n=201) | 1914 (696-2315)<br>(n=13) | 887 (410-1573)<br>(n=199) | 929 (472-1643)<br>(n=447) |
| Neutrophils, x10<sup>9</sup>/L | 8 (5.8-10.7)<br>(n=239) | 7.7 (5.4-10.2)<br>(n=47) | 7.9 (5.3-11.0)<br>(n=278) | 7.9 (5.6-10.7)<br>(n=612) |
| Lymphocytes, x10<sup>9</sup>/L | 0.7 (0.5-1.0)<br>(n=239) | 0.6 (0.4-0.9)<br>(n=47) | 0.7 (0.5-1.0)<br>(n=279) | 0.7 (0.5-1.0)<br>(n=613) |
| Platelets, x10<sup>9</sup>/L | 250 (183-323)<br>(n=346) | 273 (205-319)<br>(n=48) | 236 (177-297)<br>(n=398) | 245 (182-311)<br>(n=854) |
| Lactate, mmol/L | 1.3 (1.0-1.8)<br>(n=303) | 1.5 (1.2-2.0)<br>(n=40) | 1.4 (1.0-1.9)<br>(n=353) | 1.4 (1.0-1.9)<br>(n=754) |
| Creatinine, mg/dL | 0.8 (0.7-1.2)<br>(n=348) | 1.0 (0.6-1.4)<br>(n=48) | 0.9 (0.7-1.2)<br>(n=398) | 0.9 (0.7-1.2)<br>(n=856) |
| Bilirubin, mg/dL | 0.11 (0.08-0.15)<br>(n=327) | 0.11 (0.09-0.18)<br>(n=46) | 0.10 (0.08-0.15)<br>(n=382) | 0.11 (0.08-0.15)<br>(n=817) |

\* Percentages may not sum to 100 because of rounding. SD denotes standard deviation; APACHE, Acute Physiology and Chronic Health Evaluation; COPD, chronic obstructive pulmonary disease; IQR, interquartile range; ECMO, extracorporeal membrane oxygenation.
<sup>a</sup> Control patients include all patient randomized to control who were also eligible to be randomized to tocilizumab and/or sarilumab.
<sup>b</sup> All patients includes patient randomized in the Immune Modulation Therapy domain, including control (including where tocilizumab and sarilumab were not a randomization option), tocilizumab, sarilumab, anakinra and interferon-beta-1a.
<sup>c</sup> Data collection not approved in Canada and continental Europe. “Other” includes “declined” and “multiple”.
<sup>d</sup> The body-mass index (BMI) is the weight in kilograms divided by the square of the height in meters.
<sup>e</sup> Range 0 to 71, with higher scores indicating greater severity of illness.
<sup>f</sup> SARS-CoV2 infection was confirmed by respiratory tract polymerase chain reaction test
<sup>g</sup> Values were from the sample collected closest to randomization, up to 8 hours prior to randomization. If no samples were collected up to 8 hours prior to time of randomization, the sample collected closest to the time of randomization up to 2 hours after randomization was used (other than PaO<sub>2</sub>/FiO<sub>2</sub> which was a pre-randomization value only). Laboratory values were only added to the case report form on August 6, 2020.

In the tocilizumab group, 92% received at least one dose, 29% receiving a second dose at the discretion of the treating clinician. In the sarilumab group, 90% received the allocated drug and in the control group, 2% were given one of the immune modulating drugs outside the trial protocol.

### Primary Outcome

Median organ support-free days were 10 (interquartile range [IQR] −1, 16), 11 (IQR 0, 16) and 0 (IQR −1, 15) for tocilizumab, sarilumab and control groups, respectively (Table 2 and Figure 2). Compared with control, median adjusted odds ratios (primary model) were 1.64 (95%CrI 1.25, 2.14) for tocilizumab and 1.76 (95%CrI 1.17, 2.91) for sarilumab, yielding >99.9% and 99.5% posterior probabilities of superiority. Hospital mortality was 28.0% (98/350) for tocilizumab, 22.2% (10/45) for sarilumab and 35.8% (142/397) for control. The hospital mortality pooling both IL-6 receptor antagonists was 27.3% (108/395). Compared with control, median adjusted odds ratios for hospital survival were 1.64 (95%CrI 1.14, 2.35) for tocilizumab and 2.01 (95% CrI 1.18, 4.71) for sarilumab, yielding 99.6% and 99.5% posterior probabilities of superiority. The sensitivity analyses were consistent with the primary analysis (Tables S1 and S2). Of note, the estimates of the treatment effect for patients treated either with tocilizumab or sarilumab and corticosteroids in combination were greater than for any intervention on its own (Tables S3 and S4), suggesting benefit of using both IL6 receptor antagonists and corticosteroids together in this critically ill population.

**Table 2.** Primary and Secondary Outcomes.

| Outcome/Analysis | Tocilizumab (N=353) | Sarilumab (N=48) | Control (N=402) |
| --- | --- | --- | --- |
| **Primary Outcome, Organ support-free days (OSFDs)** |  |  |  |
| Median (IQR) | 10 (-1 to 16) | 11 (0 to 16) | 0 (-1 to 15) |
| Adjusted OR - mean (SD) | 1.65 (0.23) | 1.83 (0.44) | 1 |
| - median (95% CrI) | 1.64 (1.25 to 2.14) | 1.76 (1.17 to 2.91) | 1 |
| Probability of superiority to control, % | >99.9 | 99.5 | - |
| **Subcomponents of OSFDs** |  |  |  |
| In-hospital deaths, n (%) | 98/350 (28.0) | 10/45 (22.2) | 142/397 (35.8) |
| OSFDs in survivors, median (IQR) | 14 (7 to 17) | 15 (6.5 to 17) | 13 (4 to 17) |
| **Primary Hospital Survival,** |  |  |  |
| Adjusted OR - mean (SD) | 1.66 (0.31) | 2.25 (0.96) | 1 |
| - median (95% CrI) | 1.64 (1.14 to 2.35) | 2.01 (1.18 to 4.71) | 1 |
| Probability of superiority to control, % | 99.6 | 99.5 | - |
| **Secondary Analysis of Primary Outcome, model restricted to Immune Modulation Therapy domain patients and other closed domains** |  |  |  |
| Adjusted OR - mean (SD) | 1.68 (0.24) | 1.84 (0.44) | 1 |
| - median (95% CrI) | 1.66 (1.26 to 2.18) | 1.77 (1.18 to 2.90) | 1 |
| Probability of superiority to control, % | >99.9 | 99.6 | - |
| **Secondary Analysis of Primary Hospital Survival, model restricted to Immune Modulation Therapy domain patients and other closed domains** |  |  |  |
| Adjusted OR - mean (SD) | 1.67 (0.31) | 2.24 | 1 |
| - median (95% CrI) | 1.65 (1.15 to 2.34) | 2.00 (1.17 to 4.69) | 1 |
| Probability of superiority to control, % | 99.6 | 99.4 | - |
| **Other Secondary Outcomes** |  |  |  |
| **90-day Survival (time to event)** |  |  |  |
| Adjusted HR - mean (SD) | 1.60 (0.21) | 1.94 (0.56) | 1 |
| - median (95% CrI) | 1.59 (1.24 to 2.05) | 1.82 (1.22 to 3.38) | 1 |
| Probability of superiority to control, % | >99.9 | 99.8 | - |
| **Respiratory support-free days** |  |  |  |
| Adjusted OR - mean (SD) | 1.74 (0.25) | 2.04 (0.53) | 1 |
| - median (95% CrI) | 1.73 (1.31 to 2.27) | 1.94 (1.27 to 3.32) | 1 |
| Probability of superiority to control, % | >99.9 | 99.9 | - |
| **Cardiovascular support-free days** |  |  |  |
| Adjusted OR - mean (SD) | 1.70 (0.26) | 1.95 (0.53) | 1 |
| - median (95% CrI) | 1.68 (1.25 to 2.24) | 1.85 (1.20 to 3.30) | 1 |
| Probability of superiority to control, % | >99.9 | 99.5 | - |
| **Time to ICU discharge** |  |  |  |
| Adjusted HR - mean (SD) | 1.43 (0.13) | 1.69 (0.32) | 1 |
| - median (95% CrI) | 1.42 (1.18 to 1.70) | 1.64 (1.21 to 2.45) | 1 |
| Probability of superiority to control, % | >99.9 | 99.9 | - |
| **Time to hospital discharge** |  |  |  |
| Adjusted HR - mean (SD) | 1.42 (0.13) | 1.65 (0.31) | 1 |
| - median (95% CrI) | 1.41 (1.18 to 1.70) | 1.60 (1.17 to 2.40) | 1 |
| Probability of superiority to control, % | >99.9 | 99.8 | - |
| **WHO scale at day 14** |  |  |  |
| Adjusted OR - mean (SD) | 1.85 (0.26) | 1.91 (0.43) | 1 |
| - median (95% CrI) | 1.83 (1.40 to 2.41) | 1.86 (1.22 to 2.91) | 1 |
| Probability of superiority to control, % | >99.9 | 99.6 | - |
| **Progression to invasive mechanical ventilation, ECMO or death, restricted to those not intubated at baseline** |  |  |  |
| Free of invasive mechanical ventilation at baseline, n | 242 | 37 | 273 |
| Progression to intubation, ECMO or death, n (%) | 100/242 (41.3) | 13/37 (35.1) | 144/273 (52.7) |
| Adjusted OR - mean (SD) | 1.72 (0.33) | 1.82 (0.55) | 1 |
| - median (95% CrI) | 1.69 (1.17 to 2.42) | 1.74 (1.01 to 3.14) | 1 |
| Probability of superiority to control, % | 99.8 | 97.7 | - |
| **Serious Adverse Events** |  |  |  |
| Patients with $\geq 1$ serious adverse event, n (%) | 9/353 (2.5) | 0/48 (0.0) | 11/402 (2.7) |
| Adjusted OR - mean (SD) | 1.22 (0.55) | 2.99 (2.95) | 1 |
| - median (95% CrI) | 1.10 (0.48 to 2.58) | 2.10 (0.51 to 10.77) | 1 |
| Probability of superiority to control, % | 59.3 | 84.0 | - |

The primary analysis of organ support-free days (OSFD) and in-hospital mortality used data from all participants enrolled in the trial who met COVID-19 severe state criteria and were randomized within at least one domain (n=1928), adjusting for age, sex, time period, site, region, domain and intervention eligibility and intervention assignment.
Other analyses were restricted to participants enrolled in the Immune Modulation domain and any domains that have ceased recruitment (Corticosteroid and Covid-19 Antiviral domains) (n=1293), adjusting for age, sex, time period, site, region, domain and intervention eligibility and intervention assignment.
Definitions of outcomes are provided in Methods and the study protocol.
All models are structured such that a higher OR or HR is favorable. The WHO scale ranges from 0 (no disease) to 8 (death).
SD - standard deviation; CrI - credible interval; OR - odds ratio; HR – Hazard ratio.

**Figure 2.**
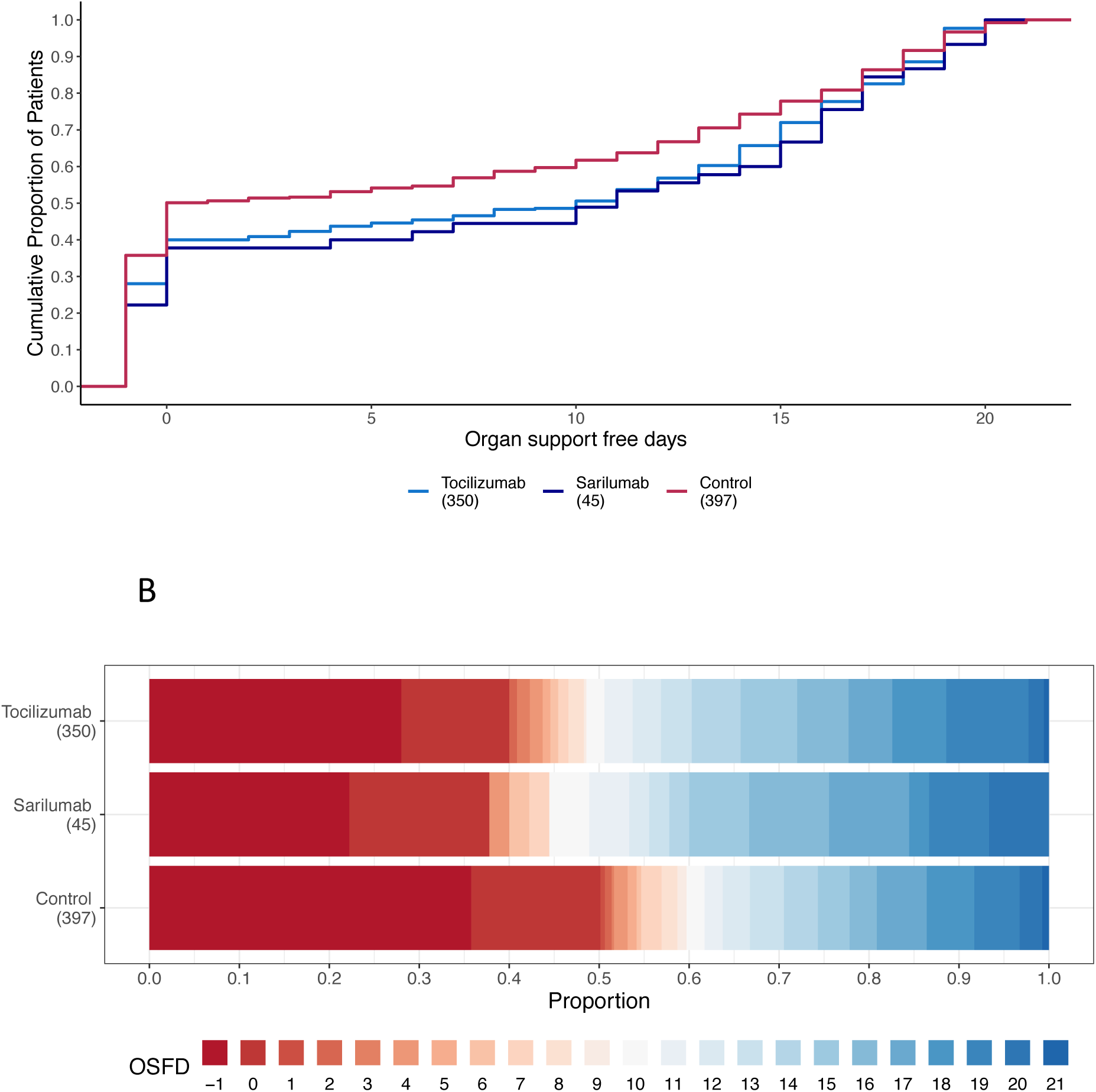
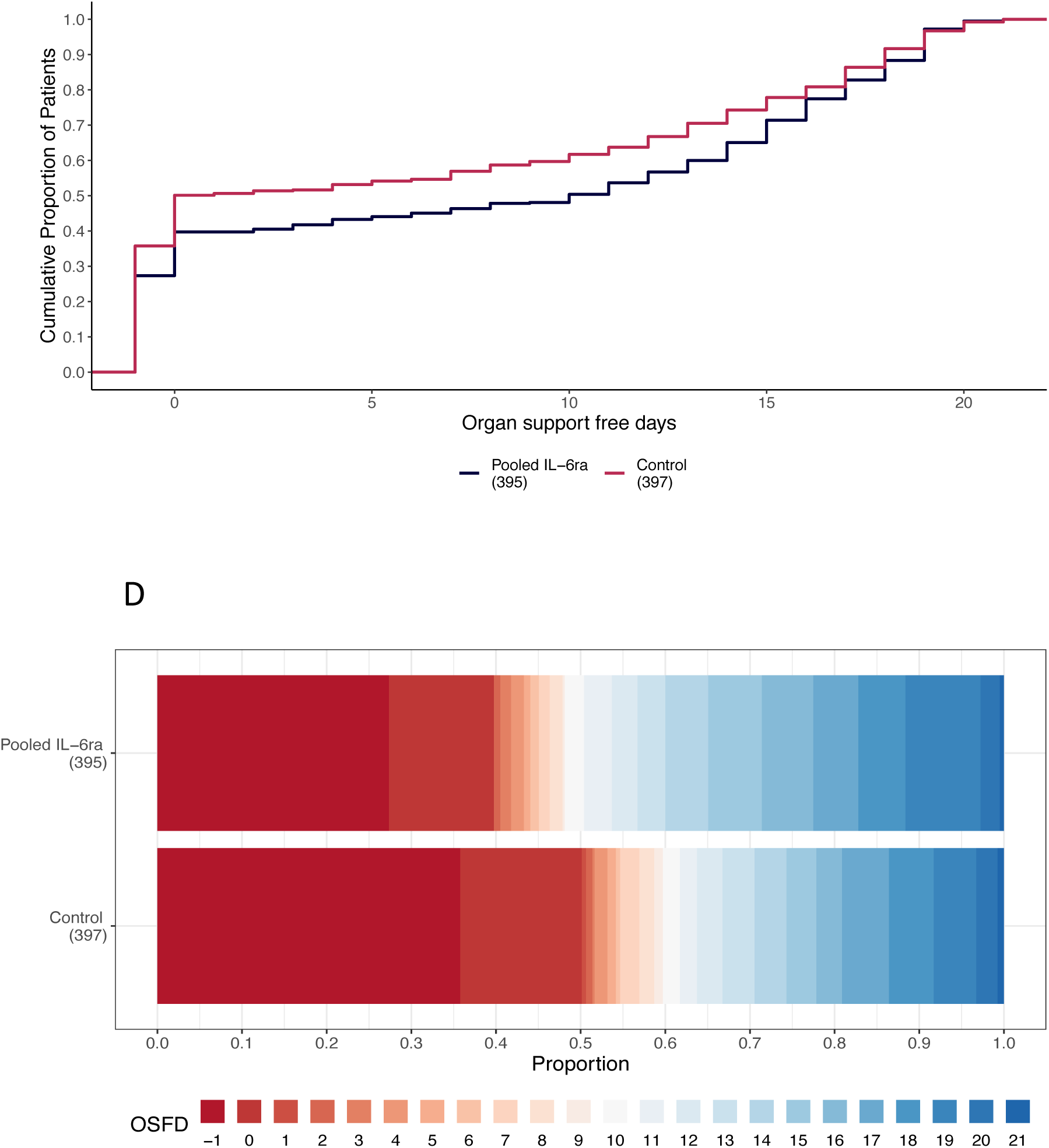
Distributions of organ support–free days. Panel A) the cumulative proportion (y-axis) for each intervention group by day (x-axis), with death listed first. Curves that rise more slowly are more favorable. Panel B) Organ support– free days as horizontally stacked proportions by intervention group. Red represents worse outcomes and blue represents better outcomes. The median adjusted odds ratios from the primary analysis, using a Bayesian cumulative logistic model, were 1.64 (95% credible interval, 1.25 to 2.14) and 1.76 (95% credible interval, 1.17 to 2.91) for the tocilizumab and sarilumab groups compared with control, yielding >99.9% and 99.5% probabilities of superiority compared with control, respectively. Panels C) and D) are similar figures with the tocilizumab and sarilumab interventions pooled together. The median adjusted odds ratio is 1.65 (95% credible interval 1.27 to 2.14) yielding >99.9% probability of superiority compared with control.

### Secondary Outcomes

The secondary outcomes are listed in Table 2 and Figure 3. Tocilizumab and sarilumab were effective across all secondary outcomes, including 90-day survival, time to ICU and hospital discharge, and improvement in the World Health Organization (WHO) ordinal scale at day 14.^16^ Similar effects were seen in all CRP subgroups (Table S1).

**Figure 3.**
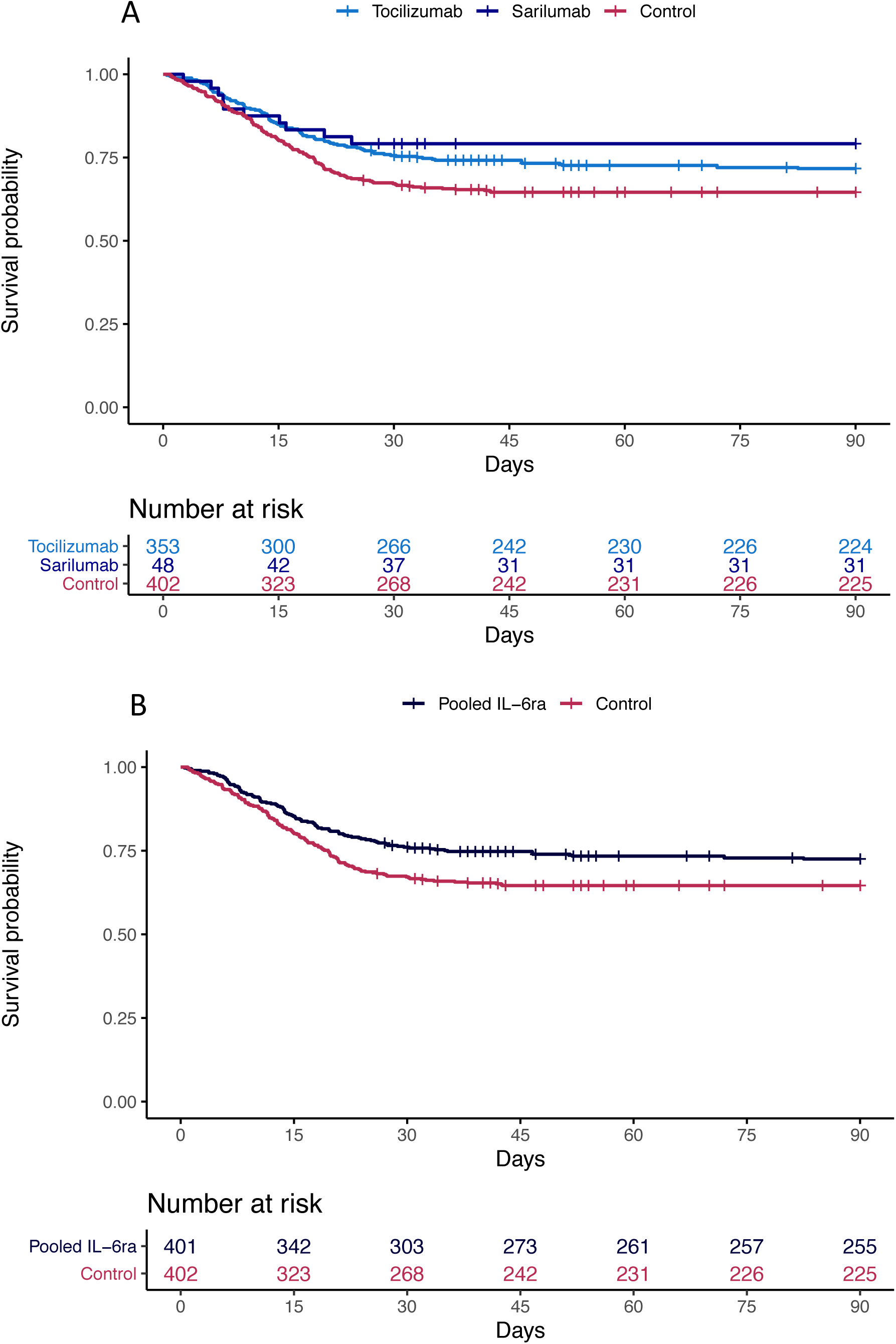
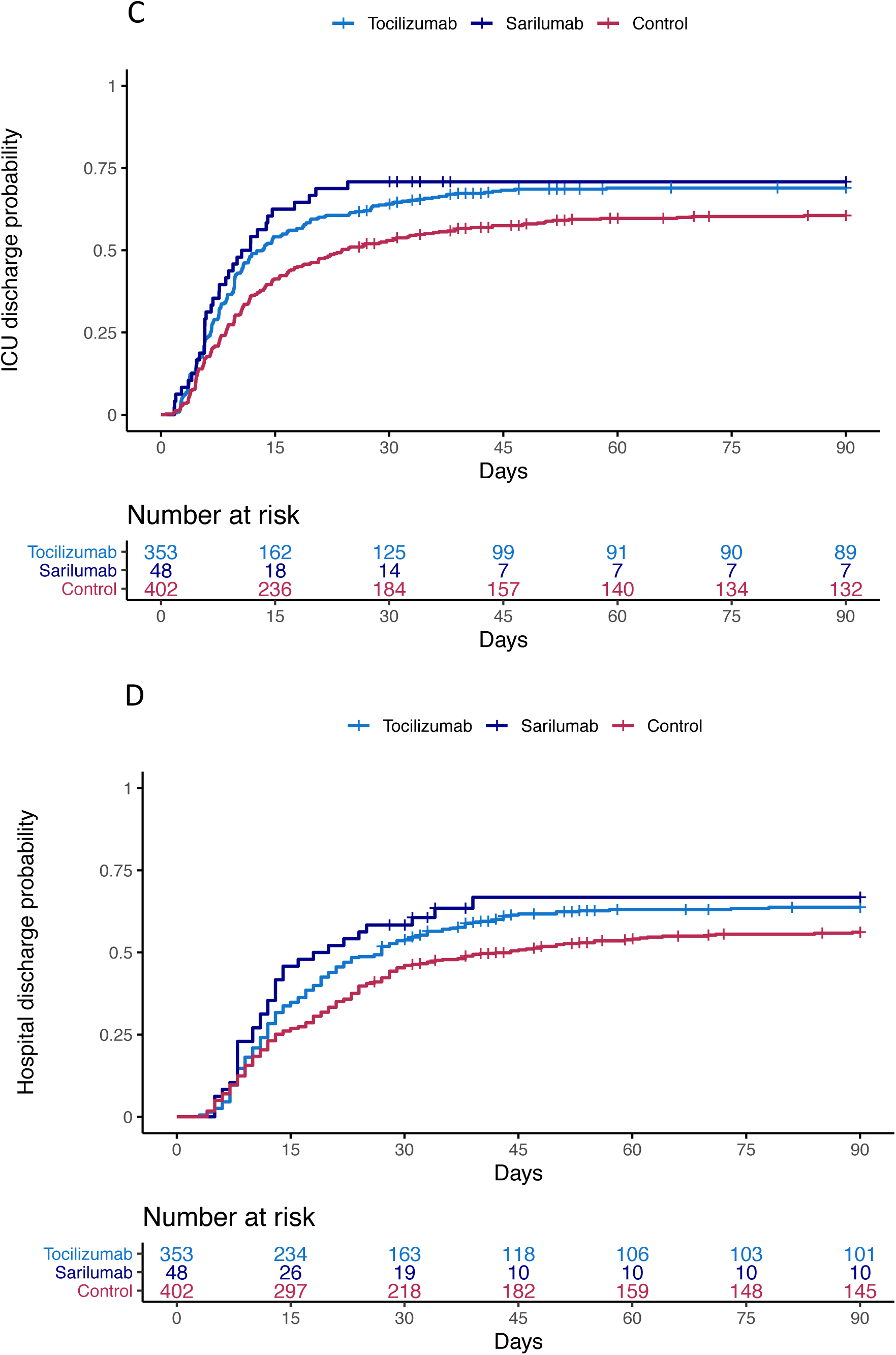
Time to event analyses. Shown are Kaplan-Meier curves for survival by individual intervention group (Panel A), survival with tocilizumab and sarilumab intervention groups pooled together (Panel B), time to intensive care unit discharge by individual intervention group (Panel C) and time to hospital discharge by individual intervention group (Panel D)

There were nine serious adverse events reported in the tocilizumab group including one secondary bacterial infection, five bleeds, two cardiac events and one deterioration in vision. There were 11 serious adverse events in the control group, four bleeds and seven thromboses; and no serious adverse events in the sarilumab group.

## Discussion

We show that in critically ill patients with Covid-19 the IL-6 receptor antagonists, tocilizumab and sarilumab, are both effective compared with current standard of care, which included corticosteroids in the majority of patients (>80%). Benefit was consistent across primary and secondary outcomes, and across subgroups and secondary analyses.

Multiple observational and ex-vivo laboratory studies have demonstrated that IL-6 is an important cytokine associated with disease severity and mortality.^17-19^ A recent genomic analysis of critically ill patients with Covid-19, demonstrated that genetic variants in the IL-6 inflammatory pathway may have a causal link to life-threatening disease.^20^ There is therefore a good rationale to support inhibiting IL-6 pathways in severe Covid-19.

Our study should be compared with other trials of IL-6 receptor antagonists in Covid-19. Many previously reported trials included less severely ill patients and excluded patients already receiving respiratory support.^6-8^ In those studies, there was no clear evidence that tocilizumab was effective at preventing disease progression, and no evidence of benefit on survival, although they may have lacked power to detect differences in patient-centered outcomes. The EMPACTA trial reported that patients treated with tocilizumab were less likely to progress to need mechanical ventilation or to die by day 28 (hazard ratio 0.56, 95%CI 0.32, 0.97), although there was no difference in overall mortality (risk difference 2.0%, 95%CI −5.2%, 7.8%).^9^ The COVACTA trial included about 38% mechanically ventilated patients. It reported no difference in clinical status or mortality at day 28, although the time to hospital discharge was shorter with tocilizumab (hazard ratio 1.35, 95%CI 1.02, 1.79).^10^ A trial of sarilumab reported no benefit in the whole population but a trend towards reduced mortality in the critically ill group.^21^ We saw both an improved time to clinical improvement as well as a reduction in mortality. It is therefore possible that the maximum benefit from IL-6 inhibition is seen in the most severely ill patients with Covid-19. However, it is important to note that in our trial, patients had to be enrolled within 24 hours after starting organ support. This may be an important factor to maximize effectiveness; treating critically ill patients early, while any developing organ dysfunction may be more reversible.

Investigators have proposed using CRP or other inflammatory markers to select patients with a hyperinflammatory state for treatment.^6,8^ We saw beneficial effects of IL-6 inhibition across all CRP subgroups in this critically ill population. Although Covid-19 has been described as producing a “cytokine storm”,^22^ recent studies have shown that systemic levels of cytokines may not be as high as seen in other causes of sepsis and ARDS.^23^ It may be that local inflammation, demonstrated by respiratory dysfunction, is a more useful indicator of which patients will benefit from IL-6 inhibition. There has been concern about administering immune modulating drugs, such as tocilizumab and sarilumab, to patients critically ill due to a novel virus infection. One consistent result across all trials to date, including our trial, is there has been no increased rates of serious adverse events reported.

REMAP-CAP’s pragmatic, international design means that our results are likely generalizable to the wider critically ill patient population with Covid-19. It, of course, has limitations. Most notably, it uses an open-label design but awareness of intervention assignment is unlikely to affect the primary outcome. Furthermore, as IL-6 inhibition is known to have a profound effect on CRP,^24,25^ even if the study drug was blinded, intervention assignment would become “revealed” rapidly after administration. As this is an early, preliminary report some data are missing including 11 outcomes. Some patients still remain in hospital and so long-term outcomes may differ from the short-term outcomes presented here. The multifactorial design also allows multiple different interventions to be evaluated simultaneously, providing more efficient results and accounting for potential treatment-by-treatment interactions. Many of these interventions continue, and their effects and possible interactions are still to be reported.

In conclusion, in critically ill adult patients with Covid-19 receiving organ support in intensive care, treatment with the IL-6 receptor antagonists, tocilizumab and sarilumab, improved outcomes, including survival.

## Data Availability

Data will be available to researchers on request subject to sponsor restrictions from 1-10-2021. Please contact

