## Supplementary Appendix for "Interleukin-6 Receptor Antagonists in Critically Ill Patients with Covid-19 – Preliminary report" (from another version of this preprint: version 1, 2021-01-07)

The REMAP-CAP Investigators

#### 1. REMAP-CAP Investigators & Collaborators

**International Trial Steering Committee:**
Farah Al-Beidh, Derek Angus, Djillali Annane, Yaseen Arabi, Abi Beane, Wilma van Bentum-Puijk, Scott Berry, Zahra Bhimani, Marc Bonten, Charlotte Bradbury, Frank Brunkhorst, Meredith Buxton, Allen Cheng, Lennie Derde, Lise Estcourt, Herman Goossens, Anthony Gordon, Cameron Green, Rashan Haniffa, Francois Lamontagne, Patrick Lawler, Edward Litton, John Marshall, Colin McArthur, Daniel McAuley, Shay McGuinness, Bryan McVerry, Stephanie Montgomery, Paul Mouncey, Srinivas Murthy, Alistair Nichol, Rachael Parke, Jane Parker, Kathryn Rowan, Marlene Santos, Christopher Seymour, Alexis Turgeon, Anne Turner, Frank van de Veerdonk, Steve Webb (Chair), Ryan Zarychanski;

**Regional Management Committees**

***Australia and New Zealand***
Yaseen Arabi, Lewis Campbell, Allen Cheng, Lennie Derde, Andrew Forbes, David Gattas, Cameron Green, Stephane Heritier, Peter Kruger, Edward Litton, Colin McArthur (Deputy Executive Director), Shay McGuinness (Chair), Alistair Nichol, Rachael Parke, Jane Parker, Sandra Peake, Jeffrey Presneill, Ian Seppelt, Tony Trapani, Anne Turner, Steve Webb (Executive Director), Paul Young

***Canadian Regional Management Committee***
Zahra Bhimani, Brian Cuthbertson, Rob Fowler, Francois Lamontagne, John Marshall (Executive Director), Venika Manoharan, Srinivas Murthy (Deputy Executive Director), Marlene Santos, Alexis Turgeon, Ryan Zarychanski

***CRIT Care Asia (CCA) Regional Management Committee***
Diptesh Aryal, Abi Beane (Chair), Arjen M Dondrop, Cameron Green, Rashan Haniffa (Executive Director), Madiha Hashmi, Issrah Jawad, Deva Jayakumar, John Marshall, Colin McArthur, Srinivas Murthy, Timo Tolppa, Vanessa Singh, Steve Webb

***European Regional Management Committee***
Farah Al-Beidh, Derek Angus, Djillali Annane, Wilma van Bentum-Puijk, Scott Berry, Marc Bonten (Executive Director and Chair), Nicole Brillinger, Frank Brunkhorst, Maurizio Cecconi, Lennie Derde, Stephan Ermann, Bruno Francois, Herman Goossens, Anthony Gordon, Cameron Green, Sebastiaan Hullegie, Rene Markgraff, Colin McArthur, Paul Mouncey, Alistair Nichol, Mathias Pletz, Pedro Povoa, Gernot Rohde, Kathryn Rowan, Steve Webb

***United States Regional Management Committee***
Brian Alexander, Derek Angus (Executive Director), Kim Basile, Meredith Buxton (Chair), Timothy Girard, Christopher Horvat, David Huang, Kelsey Linstrum, Florian Mayr, Bryan McVerry, Stephanie Montgomery, Christopher Seymour

**Regional Coordinating Centers**
***Australia, CCA region, and Saudi Arabia:*** The Australia and New Zealand Intensive Care Research Centre (ANZIC-RC), Monash University
***Canada:*** St. Michael's Hospital, Unity Health Toronto
***Europe:*** University Medical Center Utrecht (UMCU)

***Ireland:*** Irish Critical Care Clinical Trials Network, University College Dublin Research Centre, St. Vincent's Hospital
***New Zealand:*** The Medical Research Institute of New Zealand (MRINZ)
***United Kingdom:*** Intensive Care National Audit and Research Centre (ICNARC), and Imperial College London
***United States:*** Global Coalition for Adaptive Research (GCAR), and University of Pittsburgh Medical Center
**CRIT Care Asia (CCA):** NICS MORU.

__**Domain-Specific Working Groups**__

***Antibiotic and Macrolide Duration Domain-Specific Working Group***

Richard Beasley, Marc Bonten, Allen Cheng (Chair), Nick Daneman, Lennie Derde, Robert Fowler, David Gattas, Anthony Gordon, Cameron Green, Peter Kruger, Colin McArthur, Steve McGloughlin, Susan Morpeth, Srinivas Murthy, Alistair Nichol, Mathias Pletz, David Paterson, Gernot Rohde, Steve Webb

***Corticosteroid Domain-Specific Working Group***

Derek Angus (Chair), Wilma van Bentum-Puijk, Lennie Derde, Anthony Gordon, Sebastiaan Hullegie, Peter Kruger, Edward Litton, John Marshall, Colin McArthur, Srinivas Murthy, Alistair Nichol, Bala Venkatesh, Steve Webb

***Influenza Antiviral Domain-Specific Working Group***

Derek Angus, Scott Berry, Marc Bonten, Allen Cheng, Lennie Derde, Herman Goossens, Sebastiaan Hullegie, Menno de Jong, John Marshall, Colin McArthur, Srinivas Murthy (Chair), Tim Uyeki, Steve Webb

***COVID-19 Antiviral Domain-Specific Working Group***

Derek Angus, Yaseen Arabi (Chair), Kenneth Baillie, Richard Beasley, Scott Berry, Marc Bonten, Allen Cheng, Menno de Jong, Lennie Derde, Eamon Duffy, Rob Fowler, Herman Goossens, Anthony Gordon, Cameron Green, Thomas Hills, Colin McArthur, Susan Morpeth, Srinivas Murthy, Alistair Nichol, Katrina Orr, Rachael Parke, Jane Parker, Asad Patanwala, Kathryn Rowan, Steve Tong, Tim Uyeki, Frank van de Veerdonk, Steve Webb

**COVID-19 Immune Modulation Domain-Specific Working Group**

Derek Angus, Yaseen Arabi, Kenneth Baillie, Richard Beasley, Scott Berry, Marc Bonten, Frank Brunkhorst, Allen Cheng, Nichola Cooper, Olaf Cremer, Menno de Jong, Lennie Derde (Chair), Eamon Duffy, James Galea, Herman Goossens, Anthony Gordon, Cameron Green, Thomas Hills, Andrew King, Helen Leavis, John Marshall, Florian Mayr, Colin McArthur, Bryan McVerry, Susan Morpeth, Srinivas Murthy, Mihai Netea, Alistair Nichol, Kayode Ogungbenro, Katrina Orr, Jane Parker, Asad Patawala, Ville Pettilä (Deputy Chair), Emma Rademaker, Kathryn Rowan, Manoj Saxena, Christopher Seymour, Wendy Sligl, Steven Tong, Tim Uyeki, Frank van de Veerdonk, Steve Webb, Taryn Youngstein

**COVID-19 Immune Modulation -2 Domain-Specific Working Group**

Derek Angus, Scott Berry, Lennie Derde, Cameron Green, David Huang, Florian Mayr, Bryan McVerry, Stephanie Montgomery, Christopher W. Seymour (Chair), Steve Webb

***Therapeutic Anticoagulation Domain-Specific Working Group***

Derek Angus, Diptesh Aryal, Scott Berry, Shilesh Bihari, Charlotte Bradbury, Marc Carrier, Dean Fergusson, Robert Fowler, Ewan Goligher (Deputy Chair), Anthony Gordon, Christopher Horvat, David Huang, Beverley Hunt, Devachandran Jayakumar, Anand Kumar, Mike Laffan, Patrick Lawler, Sylvain Lother, Colin McArthur, Bryan McVerry, John Marshall, Saskia Middeldopr, Zoe McQuilten, Matthew Neal, Alistair Nichol, Christopher Seymour, Roger Schutgens, Simon Stanworth, Alexis Turgeon, Steve Webb, Ryan Zarychanski (Chair)

***Vitamin C Domain-Specific Working Group***
Neill Adhikari (Chair), Derek Angus, Djillali Annane, Matthew Anstey, Yaseen Arabi, Scott Berry, Emily Brant, Angelique de Man, Lennie Derde, Anthony Gordon, Cameron Green, David Huang, Francois Lamonagne (Chair), Edward Litton, John Marshall, Marie-Helene Masse, Colin McArthur, Shay McGuinness, Paul Mouncey, Srinivas Murthy, Rachael Parke, Alistair Nichol, Tony Trapani, Andrew Udy, Steve Webb

***COVID-19 Immunoglobulin Domain-Specific Working Group***
Derek Angus, Donald Arnold, Phillipe Begin, Scott Berry, Richard Charlewood, Michael Chasse, Mark Coyne, Jamie Cooper, James Daly, Lise Estcourt (Chair, UK lead), Dean Fergusson, Anthony Gordon, Iain Gosbell, Heli Harvala-Simmonds, Tom Hills (New Zealand lead), Christopher Horvat, David Huang, Sheila MacLennan, John Marshall, Colin McArthur (New Zealand lead), Bryan McVerry (USA lead), David Menon, Susan Morpeth, Paul Mouncey, Srinivas Murthy, John McDyer, Zoe McQuilten (Australia lead), Alistair Nichol (Ireland lead), Nicole Pridee, David Roberts, Kathryn Rowan, Christopher Seymour, Manu Shankar-Hari (UK lead), Helen Thomas, Alan Tinmouth, Darrell Triulzi, Alexis Turgeon (Canada lead), Tim Walsh, Steve Webb, Erica Wood, Ryan Zarychanski (Canada lead)

***Simvastatin Domain-Specific Working Group***
Derek Angus, Yaseen Arabi, Abi Beane, Carolyn Calfee, Anthony Gordon, Cameron Green, Rashan Haniffa, Deva Jayakumar, Peter Kruger, Patrick Lawler, Edward Litton, Colin McArthur, Daniel McAuley (Chair), Bryan McVerry, Matthew Neal, Alistair Nichol, Cecilia O'Kane, Murali Shyamsundar, Pratik Sinha, Taylor Thompson, Steve Webb, Ian Young

***Antiplatelet Domain-Specific Working Group***
Derek Angus, Scott Berry, Shailesh Bihari, Charlotte Bradbury (Chair), Marc Carrier, Timothy Girard, Ewan Goligher, Anthony Gordon, Ghady Haidar, Christopher Horvat, David Huang, Beverley Hunt, Anand Kumar, Patrick Lawler, Patrick Lawless, Colin McArthur, Bryan McVerry, John Marshall, Zoe McQuilten, Matthew Neal, Alistair Nichol, Christopher Seymour, Simon Stanworth, Steve Webb, Alexandra Weissman, Ryan Zarychanski

***Mechanical Ventilation Domain***
Derek Angus, Wilma van Bentum-Puijk, Lewis Campbell, Lennie Derde, Niall Ferguson, Timothy Girard, Ewan Goligher, Anthony Gordon, Cameron Green, Carol Hodgson, Peter Kruger, John Laffey, Edward Litton, John Marshall, Colin McArthur, Daniel~~ny~~ McAuley, Shay McGuinness, John Laffey, Neil Orford, Kathryn Rowan, Ary Neto, Steve Webb

***ACE-2 RAS Domain***
Rebecca Baron, Lennie Derde, Slava Epelman, Claudia Frankfurter, David Gattas, Frank Gommans, Anthony Gordon, Rashan Haniffa, David Huang, Edy Kim, Francois Lamontagne, Patrick Lawler (Chair), David Leaf, John Marshall, Colin McArthur, Bryan McVerry, Daniel McAuley, Muthiah Vaduganathan, Roland van Kimmenade, Frank van de Veerdonk, Steve Webb

**Statistical Analysis Committee**
Michelle Detry, PhD, Mark Fitzgerald, PhD, Roger Lewis, MD, PhD (Chair), Anna McGlothlin, PhD, Ashish Sanil, PhD, Christina Saunders, PhD

**Statistical Design Team**
Lindsay Berry, PhD, Scott Berry, PhD, Elizabeth Lorenzi, PhD

**Project Management**
***Australia and Saudi Arabia:*** Jane Parker, Vanessa Singh, Claire Zammit
***Canada:*** Zahra Bhimani, Marlene Santos
***CCA:*** Abi Beane, Rashan Haniffa, Timo Tolppa
***Europe:*** Wilma van Bentum Puijk, Wietske Bouwman, Radhika Ganpat, Erika Groenveld, Denise van Hout, Yara Mangindaan, Clementina Okundaye, Lorraine Parker, Svenja Peters, Ilse Rietveld, Linda Rikkert, Kik Raymakers, Irma Scheepstra-Beukers, Albertine Smit,
***Germany:*** Nicole Brillinger, Rene Markgraf
***Global:*** Cameron Green
***Ireland:*** Kate Ainscough, Kathy Brickell, Peter Doran
***New Zealand:*** Anne Turner
***United Kingdom:*** Farah Al-Beidh, Aisha Anjum, Janis-Best Lane, Elizbeth Fagbodun, Lorna Miller, Paul Mouncey, Karen Parry-Billings, Sam Peters, Alvin Richards-Belle, Michelle Saull, Stefan Sprinkmoller, Daisy Wiley
***United States of America:*** Kim Basile, Meredith Buxton, Kelsey Linstrum, Stephanie Montgomery

**Data and Safety Monitoring Board**
Julian Bion, Jason Connor, (Deputy Chair), Simon Gates, Victoria Manax (Chair), Tom van der Poll, John Reynolds

**Database Providers**
***Research Online:***
Marloes van Beurden, Evelien Effelaar, Joost Schotsman,
***Spinnaker Software:***
Craig Boyd, Cain Harland, Audrey Shearer, Jess Wren
***University of Pittsburgh Medical Center***
Giles Clermont, William Garrard, Christopher Horvat, Kyle Kalchthaler, Andrew King, Daniel Ricketts, Salim Malakoutis, Oscar Marroquin, Edvin Music, Kevin Quinn
**NICS MORU: on behalf of CCA;**
Udara Attanayaka, Abi Beane, Sri Darshana, Rashan Haniffa, Pramodya Ishani, Issrah Jawad, Upulee Pabasara, Timo Tolppa, Ishara Udayanga.

**Clinical Trials Groups**
The REMAP-CAP platform is supported by the Australian and New Zealand Intensive Care Society Clinical Trials Group, the Canadian Critical Care Clinical Trials Group, the Irish Critical Care Clinical Trials Network, the UK Critical Care Research Group and the International Forum of Acute Care Trialists.

REMAP-CAP was supported in the UK by the NIHR Clinical Research Network and we acknowledge the contribution of Kate Gilmour, BSc (Hons), Karen Pearson, MSc, Chris Siewerski, MSc, Sally-Anne

Hurford, MSc, Emma Marsh, FdSC, Debbie Campbell, Penny Williams, MSc, Kim Shirley, LLB Hons, Meg Logan, NVQ, Jane Hanson, Becky Dilley, BSc, Louise Phillips, CIM, Anne Oliver, MSc, Mihaela Sutu, MSc, Sheenagh Murphy, PGDip, Latha Aravindan, PhD, Joanne Collins, MRes, Holly Monaghan, Adam Unsworth, NVQ, Seonaid Beddows, MSc, Laura Ann Dawson, LLM, Sarah Dyas, Adeeba Asghar, MSc, Kate Donaldson, BA, Tabitha Skinner, BSc, Nhlanhla Mguni, BSc (Hons), Natasha Muzengi, BSc, Ji Luo, PhD, Joanna O'Reilly, BSc (Hons), Chris Levett, MSc, Alison Potter, David Porter, PhD, Teresa Lockett, MSc, Jazz Bartholomew, MSc, Clare Rook, MSc, Rebecca McKay, Hannah Williams, MSc, Alistair Hall, FRCP, Hilary Campbell, BSc (Hons), Holly Speight, BA, Sandra Halden, Susan Harrison, Mobeena Naz, Kaatje Lomme, MA, Paula Sharratt, MSc, Johnathan Sheffield, FRCP, William Van't Hoff, FRCPCH, James D Williamson, PhD, Alex Barnard, BSc (Hons), Catherine Birch, BA, Morwenna Brend, PhD, Emma Chambers, PhD, Sarah Crawshaw, PhD, Chelsea Drake, BSc, Hayley Duckles-Leech, PhD, Justin Graham, BSc (Hons), Heather Harper, BSc (Hons), Stephen Lock, PGCert, Nicola McMillan, PhD, Cliodhna O'Flaherty, Eleanor OKell, PhD, Amber Hayes, Sally Sam, BSc, Heather Slade, MSc, Susan Walker, PhD, Karen Wilding, MRes, Jayne Goodwin, Helen Hodgson, Yvette Ellis, Dawn Williamson, Madeleine Bayne, MSc, Shane Jackson, Rahim Byrne, Sonia McKenna, Alsion Clinton, NIHR Urgent Public Health Group: https://www.nihr.ac.uk/documents/urgent-public-health-group-members/24638#Members

REMAP-CAP was supported in France by the CRICS-TRIGGERSEP network

REMAP-CAP was supported in Ireland by the Irish Critical Care Clinical Trials Network and we acknowledge the contribution of Kate Ainscough, Kathy Brickell and Peter Doran.

REMAP-CAP was supported in the Netherlands by the Research Collaboration Critical Care the Netherlands (RCC-Net).

Site Investigators and Research Coordinators

***Australia:***

*The Alfred Hospital*: Andrew Udy, PhD, Phoebe McCracken, MPH, Meredith Young, MPH, Jasmin Board, MPH, Emma Martin, BPharmSc;

*Ballarat Health Services*: Khaled El-Khawas, FCICM, Angus Richardson, FCICM, Dianne Hill, BN GradCert(CritCare), Robert J Commons, PhD, Hussam Abdelkharim, FCICM;

*Bendigo Hospital*: Cameron Knott, MBBS(Hons) GDipClinUS MClinEd FCICM FRACP, Julie Smith, GDipN(CritCare), Catherine Boschert, PGDip;

*Caboolture Hospital*: Julia Affleck, MBiotech, Yogesh Apte, FCICM, Umesh Subbanna, FCICM, Roland Bartholdy, FCICM, Thuy Frakking, PhD;

*Campbelltown Hospital*: Karuna Keat, MB BS(Hons), Deepak Bhonagiri, MD, Ritesh Sanghavi, MBBS, Jodie Nema, B.App.Sc.(Ag), Megan Ford, BSc;

*Canberra Hospital*: Harshel G. Parikh, FCICM, Bronwyn Avard, MLMEd, Mary Nourse, PGCert(CritCare);

*Concord Repatriation General Hospital*: Winston Cheung, MBChB, Mark Kol, MBBS, Helen Wong, RN, Asim Shah, MBChB, Atul Wagh, MBBS;

*Eastern Health (Box Hill, Maroondah & Angliss Hospitals)*: Joanna Simpson, FCICM FANZCA, Graeme Duke, FCICM, Peter Chan, FCICM, Brittney Carter, PGCert(CritCare), Stephanie Hunter, MN;

*Flinders Medical Centre*: Shailesh Bihari, PhD, Russell D Laver, FCICM, Tapaswi Shrestha, MN, Xia Jin, MN;

*Fiona Stanley Hospital*: Edward Litton, FCICM, Adrian Regli, FCICM, Susan Pellicano, PGCert, Annamaria Palermo, BA, Ege Eroglu, BSc(Hons);

*Footscray Hospital*: Craig French, MBBS, Samantha Bates, RN GDipCritCare, Miriam Towns, MPH, Yang Yang, MBBS, Forbes McGain, PhD;

*Gold Coast University Hospital*: James McCullough, FCICM, Mandy Tallott, MN;

*John Hunter Hospital*: Nikhil Kumar, MBBS, Rakshit Panwar, FCICM, Gail Brinkerhoff, PGCert, Cassandra Koppen, BPharm, Federica Cazzola, MBBS;

*Launceston General Hospital*: Matthew Brain, FCICM, FRACP, DDU; Sarah Mineall, MN;

*Lyell McEwin Hospital*: Roy Fischer, FCICM, Vishwanath Biradar, FCICM, Natalie Soar, BSN;

*Logan Hospital*: Hayden White, PhD, Kristen Estensen, MBBS, Lynette Morrison, PGCert(CritCare), Joanne Sutton, RN, Melanie Cooper, PGCert(CritCare);

*Monash Health (Monash Medical Centre, Dandenong Hospital & Casey Hospital):* Yahya Shehabi, PhD, Wisam Al-Bassam, MBChB, Amanda Hulley, BSc; Umesh Kadam, MSc, Kushaharan Sathianathan, MSc;

*Nepean Hospital*: Ian Seppelt, PhD, Christina Whitehead, MN, Julie Lowrey, BN, Rebecca Gresham, BN, Kristy Masters, BN;

*Princess Alexandra Hospital*: Peter Kruger, PhD, James Walsham, FCICM, Mr Jason Meyer, BN, Meg Harward, MN, Ellen Venz, PGCert(CritCare);

*The Prince Charles Hospital*: Kara Brady, MPharm, Cassandra Vale, GDipClinPharm, Kiran Shekar, PhD, Jayshree Lavana, FCICM, Dinesh Parmar, FCICM;

*The Queen Elizabeth Hospital*: Sandra Peake, PhD, Patricia Williams, BN, Catherine Kurenda, RN;

*Rockhampton Hospital*: Helen Miles, MBBS MPH FCICM, Antony Attokaran, MBBS FCICM FRACP;

*Royal Adelaide Hospital*: Samuel Gluck, MD, Stephanie O'Connor, MNSc, Marianne Chapman, PhD, Kathleen Glasby, CCRN;

*Royal Darwin Hospital*: Lewis Campbell, FCICM, Kirsty Smyth, PGCert(CritCare), Margaret Phillips, MN;

*Royal Melbourne Hospital*: Jeffrey Presneill, PhD, Deborah Barge, CCRN, Kathleen Byrne, MNSc, Alana Driscoll GDipN(CritCare), Louise Fortune, MNSc;

*Royal North Shore Hospital*: Pierre Janin, MD, Elizabeth Yarad, MN, Frances Bass, MSc, Naomi Hammond, PhD, Anne O'Connor, RN;

*Royal Perth Hospital*: Sharon Waterson, PGCert(CritCare), Steve Webb, PhD, Robert McNamara, BMBS;

*Royal Prince Alfred Hospital*: David Gattas, MMed(ClinEpid), Heidi Buhr, MScMed(ClinEpid), Jennifer Coles, GDipN(CritCare);

*Sir Charles Gardiner Hospital*: Sacha Schweikert, FCICM, Bradley Wibrow, FCICM, Matthew Anstey, FCICM, Rashmi Rauniyar, MPH;

*St George Hospital*: Kush Deshpande, FCICM, Pam Konecny, FRACP, Jennene Miller, BN, Adeline Kintono, BN, Raymond Tung B.Pharm, MPS, MSHP

*St. John of God Midland Public and Private Hospitals*: Ed Fysh, PhD, Ashlish Dawda, MD, Bhaumik Mevavala, MHA;

*St. John of God Hospital, Murdoch*: Annamaria Palermo, BA, Adrian Regli, FCICM, Bart De Keulenaer, FCICM;

*St. John of God Hospital, Subiaco*: Ed Litton, PhD, Janet Ferrier, BSc;

*St. Vincent's Hospital (NSW)*: Priya Nair, PhD, Hergen Buscher, FCICM, Claire Reynolds, MClinNurse, Sally Newman, PGCert(CritCare);

*St. Vincent's Hospital (VIC)*: John Santamaria, MDBS, Leanne Barbazza, PGCert(CritCare), Jennifer Homes, PGCert(CritCare), Roger Smith, MPH;

*Sunshine Coast University Hospital*: Peter Garrett, FCICM, Lauren Murray, MSc, Jane Brailsford, PGCert(CritCare), Loretta Forbes, PGCert(CritCare), Teena Maguire, BA;

*Sunshine Hospital*: Craig French, MBBS, Gerard Fennessy, MBBS, John Mulder, MBBS, Rebecca Morgan, RN PGCert(CritCare), Rebecca McEldrew, RN PGCert(CritCare);
*The Sutherland Hospital*: Anas Naeem, MClinEd, Laura Fagan, GDipN(CritCare), Emily Ryan, PGCert(CritCare);
*Toowoomba Hospital*: Vasanth Mariappa, FCICM, Judith Smith, PGCert;
*University Hospital Geelong*: Scott Simpson, MBBS, Matthew Maiden, PhD, Allison Bone, GDipN(CritCare), , Michelle Horton, MN, Tania Salerno, PGCert;
*Wollongong Hospital*: Martin Sterba, PhD, Wenli Geng, MN;

***Belgium:***
*Ghent University Hospital*: Pieter Depuydt, PhD, Jan De Waele, PhD, Liesbet De Bus, PhD, Jan Fierens, MD, Stephanie Bracke, BSc, Joris Vermassen, MD, Daisy Vermeiren, BSc;

***Canada*:**
*Brantford General Hospital:* Brenda Reeve, MD, William Dechert, MSc;
Centre de recherche de l'Institut universitaire de cardiologie et de pneumologie de Québec: Francois Lellouche, Patricia Lizotte
*Centre Hospitalier de l'Universite de Montreal:* Michaël Chassé, PhD, François Martin Carrier, MSc, Dounia Boumahni, BSc, Fatna Benettaib, MSc., Ali Ghamraoui, BSc;
*CHU de Québec – Université Laval*: Alexis Turgeon, MSc, David Bellemare, BSc, Ève Cloutier, Rana Daher, François Lauzier, MSc, Charles Francoeur, MSc;
*Centre Hospitalier Universitaire de Sherbrooke:* François Lamontagne, MD, Frédérick D'Aragon, MD, Elaine Carbonneau, BACC, Julie Leblond, BACC;
Grace Hospital: Gloria Vazquez-Grande, Nicole Marten
Grand River Hospital (Kitchener): Theresa Liu, Atif Siddiqui
*Health Sciences Centre, Winnipeg:* Ryan Zarychanski, MD, Gloria Vazquez-Grande, MD, Nicole Marten, RN, Maggie Wilson, MSc;
*Hôpital du Sacré Coeur de Montréal:* Martin Albert, MD, Karim Serri, MD, Alexandros Cavayas, MD, Mathilde Duplaix, MSc, Virginie Williams, PhD;
*Juravinski Hospital:* Bram Rochwerg, MD, Tim Karachi, MD, Simon Oczkowski, MD, John Centofanti, MD, Tina Millen, RRT
McGill University Health Centre: Kosar Khwaja, Josie Campisi
*Niagara Health (St. Catherine's Hospital):* Erick Duan, MD, Jennifer Tsang, MD, Lisa Patterson, BA;
*Regina General Hospital*: Eric Sy, Chiraag Gupta, Sandy SHA Kassir
*Royal Alexandra Hospital*: Demetrios Kutsogiannis, Patricia Thompson
*Sunnybrook Health Sciences Centre*: Rob Fowler, Neill Adhikari, Maneesha Kamra, Nicole Marinoff
*St. Boniface General Hospital*: Ryan Zarychanski, Nicole Marten
*St. Joseph's Healthcare Hamilton*: Deborah Cook, Frances Clarke
*St. Mary's General Hospital (Kitchener)*: Rebecca Kruisselbrink, Atif Siddiqui
*St. Michael's Hospital*: John Marshall, MD, Laurent Brochard, MD, Karen Burns, MD, Gyan Sandhu, RN, Imrana Khalid, MD;
*The Ottawa Hospital:* Shane English, MSc, Irene Watpool, BScN, Rebecca Porteous, BSN, Sydney Miezitis, BSc, Lauralyn McIntyre, MSc;
*University Health Network:* Elizabeth Wilcox, Lorenzo del Sorbo, Hesham Abdelhady, Tina Romagnuolo
*University of Alberta:* Wendy Sligl, Nadia Baig
*William Osler Health System:* Alexandra Binnie, MD, Elizabeth Powell, MD, Alexandra McMillan, MD, Tracy Luk, MD, Noah Aref, MSc

**CCA:**
India Apollo Speciality Hospital - OMR, Chennai: Devachandran Jayakumar, MD, Suresh Babu BSc; Apollo Main Hospital, Chennai: C Vignesh, MD, Augustian James BSc.
Apollo Speciality Vanagaram, Vanagaram, Chennai; R Ebenezer MD, S Krishnamurthy MD, Lakshmi Ranganathan PhD, Manisha, MD
Nepal Grande International Hospital : Sushil Khanal, MD, Sameena Amatya, RN; HAMS Hospital: Hem Raj Paneru, MD, Sabin Koirala, MD, Pratibha Paudel ,RN; Nepal Mediciti Hospital: Diptesh Aryal,MD, Kanchan Koirala,RN, Namrata Rai,RN, Subekshya Luitel, BSc; Tribhuvan University Teaching Hospital : Hem Raj Paneru,MD, Binita Bhattarai, RN;
Pakistan (CCA):
Ziauddin Hospital Clifton Campus: Madiha Hashmi, MD; Ashok Panjwani, MD; Zulfiqar Ali Umrani, Shoaib Siddiq; Mohiuddin Shaikh; National Institute of Cardiovascular Diseases Pakistan: Nawal Salahuddin, MD, Sobia Masood;

***Croatia:***
*General Hospital Pozega*: Zdravko Andric, Sabina Cviljevic, Renata Đimoti, Marija Zapalac, Gordan *Mirković;*
*University Hospital of Infectious Diseases "Dr Fran Milhajevid"*: Bruno Baršić, Marko Kutleša, Viktor Kotarski;
*University Hospital of Zagreb*: Ana Vujaklija Brajković, Jakša Babel, Helena Sever, Lidija Dragija, Ira Kušan;

***Finland:***
*Helsinki University Hospital*: Suvi Vaara, PhD, Leena Pettilä, Jonna Heinonen, Ville Pettilä, PhD;
Tampere University Hospital: Anne Kuitunen, PhD, Sari Karlsson, PhD, Annukka Vahtera, PhD, Heikki Kiiski, PhD, Sanna Ristimäki***;***

***France:***
*Ambroise Pare Hospital*: Amine Azaiz, Cyril Charron, MD, Mathieu Godement, MD, Guillaume Geri, Antoine Vieillard-Baron;
*Centre Hospitalier de Melun*: Franck Pourcine, Mehran Monchi;
*Centre Hospitalier Simone Veil, Beauvais*: David Luis, MD, Romain Mercier, MD, Anne Sagnier, MD, Nathalie Verrier, MD, Cecile Caplin, MD, Jack Richecoeu, MD, Daniele Combaux, MD;
*Centre Hospitalier Sud Essonne*: Shidasp Siami, PhD, Christelle Aparicio, Sarah Vautier, Asma Jeblaoui, Delphine Lemaire-Brunel;
*Centre Hospitalier Tenon*: Muriel Fartoukh, MD, Laura Courtin, Vincent Labbe, MD , Guillaume Voiriot, MD, Sara Nesrine Salhi;
*Centre Hospitalier Victor Dupouy*: Gaetan Plantefeve, MD, Cécile Leparco, RN, Damien Contou, MD ;
CHR d'Orleans: Grégoire Muller, MD, Mai-Anh Nay, MD, Toufik Kamel, MD, Dalila Benzekri, MD, Sophie Jacquier, MD, Isabelle Runge, MD, Armelle Mathonnet, MD, François Barbier, MD, Anne Bretagnol, MD;
*CHRU Tours Hopital Bretonneau*: Emmanuelle Mercier, MD, Delphine Chartier, Charlotte Salmon, MD, Pierre-François Dequin, PhD, Denis GAROT, MD;
*CHU Dupuytren, Limoges;*
*Hôpital Civil, Hôpitaux Universitaires de Strasbourg;*
*Hôpital de Hautepierre, Hôpitaux Universitaires de Strasbourg*: Francis Schneider, PhD, Vincent Castelain, PhD, Guillaume Morel, MD, Sylvie L'Hotellier, MSc;

Hospital Nord Franche-Comté: Julio Badie, MD, Fernando Daniel Berdaguer, MD, Sylvain Malfroy, MD, Chaouki Mezher, MD, Charlotte Bourgoin, MSc, Guy Moneger, MD, Elodie Bouvier, MSc;
Lariboisière Hospital: Bruno Megarbane, PhD, Sebastian Voicu, PhD, Nicolas Deye, PhD, Isabelle Malissin, MD, Laetitia Sutterlin, MD, Aymen Mrad, MD, Adrien Pépin Lehalleur, MD, Giulia Naim, MD, Philippe Nguyen, MD, Jean-Michel Ekhérian, MD, Yvonnick Boué, MD, Georgios Sidéris, PhD, Dominique Vodovar, PhD, Emmanuelle Guérin, MD, Caroline Grant, MD;
Le Mans Hospital: Christophe Guitton, PhD, Cédric Darreau, MD, Mickaël Landais, MD, Nicolas Chudeau, MD, Alain Robert, PhD, Patrice Tirot, MD, Jean Christophe Callahan, MD, Marjorie Saint Martin, MD, Charlène Le Moal, MD, Rémy Marnai, MD, Marie Hélène Leroyer;
Raymond Poincaré Hospital: Djillali Annane, MD, Pierre Moine, MD, Nicholas Heming, MD, Virginie Maxime, MD, Isabelle Bossard, MSc, Tiphaine Barbarin Nicholier, MD, Bernard Clair, MD, David Orlikowski, MD, Rania Bounab, MD, Lilia Abdeladim, MD;
*Vendee Hospital*: Gwenhael Colin, MD, Vanessa Zinzoni, Natacha Maquigneau, Matthieu Henri-Lagarrigue, MD, Caroline Pouplet, MD;

***Germany:***
*Carl-Thiem-Klinikum Cottbus gGmbH*: Jens Soukup, PD, Richard Wetzold, Madlen Löbel, Dr. Ing, Lisa Starke, Patrick Grimm;
*Charité - Universitätsmedizin Berlin*: André Finn, MD, Gabriele Kreß, Uwe Hoff, MD, Carl Friedrich Hinrichs, MD, Jens Nee, MD;
*Jena University Hospital*: Mathias W. Pletz, PhD, Stefan Hagel, PhD, Juliane Ankert, MSc, Steffi Kolanos, BSc, Frank Bloos, PhD;
*Klinikum Dortmund gGmbH*: Daniela Nickoleit-Bitzenberger, MD , Bernhard Schaaf, MD, Werner Meermeier, MD, Katharina Prebeg, Harun Said Azzaui, Martin Hower, Klaus-Gerd Brieger, Corinna Elender, Timo Sabelhaus, Ansgar Riepe, Ceren Akamp, Julius Kremling, Daniela Klein, Elke Landsiedel-Mechenbier;
*University Hospital of Leipzig*: Sirak Petros, MD, Kevin Kunz, MD, Bianka Schütze, BSc;
*Universitätsklinikum Hamburg-Eppendorf*: Stefan Kluge, MD, Axel Nierhaus, MD, Dominik Jarczak, MD, Kevin Roedl, MD;
*University Hospital of Frankfurt*: Gernot Gerhard Ulrich Rohde, MD, Achim Grünewaldt, MD, Jörg Bojunga, MD;
*University Hospital of Wuerzburg*: Dirk Weismann, MD, Anna Frey, MD; Maria Drayss, MD, M.E. Goebeler, MEG, Thomas Flor, Gertrud Fragner, Nadine Wahl, Juliane Totzke, Cyrus Sayehli, MD;
*Vivantes Klinikum Neukölln*: Lorenz Reill, Michael Distler, MD, Astrid Maselli;

***Hungary:***
*Almási Balogh Pál Hospital, Ózd*: János Bélteczki, István Magyar, Ágnes Fazekas, Sándor Kovács, Viktória Szőke;
*Jósa András County Hospital, Nyíregyháza*: Gábor Szigligeti, János Leszkoven;

***Ireland:***
*Beacon Hospital Dublin*: Daniel Collins, MRCPI, Kathy Brickell, RGN, Liadain Reid, MPH, Michelle Smyth, PgDip, Patrick Breen, MB FJFICMI, Sandra Spain, RGN;
*Beaumont Hospital*: Gerard Curley; PhD, Natalie McEvoy, MSc, Pierce Geoghegan, MB, Jennifer Clarke, MB;

*Galway University Hospitals*: John Laffey, MD, Bairbre McNicholas, PhD, Michael Scully, MD, Siobhan Casey, RN, Maeve Kernan, RN, Aoife Brennan, PhD, Ritika Rangan, PhD, Riona Tully, MB, Sarah Corbett, MB, Aine McCarthy, MB, Oscar Duffy, MB, David Burke, MB;

*St Vincent's University Hospital, Dublin*: Alistair Nichol, PhD, Kathy Brickell, RGN, Michelle Smyth, PGDip, Leanne Hayes, PhD, Liadain Reid, MPH, Lorna Murphy, RGN, Andy Neill, MB, Bryan Reidy, MSc, Michael O'Dwyer, PhD, Donal Ryan, MD, Kate Ainscough, PhD;

***Netherlands:***

*Canisius Wilhelmina Ziekenhuis*: Oscar Hoiting, MD, Marco Peters, MD, Els Rengers, MD, Mirjam Evers, RN, Anton Prinssen, RN;

*Deventer Hospital*: Huub L.A. van den Oever, MD, Arriette Kruisdijk-Gerritsen, CCRN;

*Jeroen Bosch Ziekenhuis*: Koen Simons, PhD, Tamara van Zuylen, RN, Angela Bouman, RN;

*Meander Medisch Centrum*: Laura van Gulik, PhD;

*Radboud University Medical Center Nijmegen*: Jeroen Schouten, PhD, Peter Pickkers, PhD, Noortje Roovers, BSc, Margreet Klop-Riehl, BSc, Hetty van der Eng, BSc;

*UMC Leiden*: Evert de Jonge, PhD, Jeanette Wigbers, RN, Michael del Prado, RN;

*UMC Utrecht*: Marc Bonten, PhD, Olaf Cremer, PhD, Lennie Derde, PhD, Diederik van Dijk, PhD, Emma Rademaker, MD, Jelle Haitsma Mulier, MD, Anna Linda Peters, PhD, Birgit Romberg, MD;

Ziekenhuis Gelderse Vallei: Sjoerd van Bree, MD, Marianne Bouw-Ruiter, RN, Barbara Festen, MD, Fiona van Gelder, MD, Mark van Iperen, MD, Margreet Osinga, RN, Roel Schellaars, MD, Dave Tjan, MD, Ruben van der Wekken, MD, Max Melchers, MD, Arthur van Zanten, MD;

***New Zealand***:

*Auckland City Hospital, Cardiothoracic and Vascular ICU:* Shay McGuinness, MBChB, Rachael Parke, PhD, Eileen G~~u~~ilder, MA, Magdalena Butler, RN, Keri-Anne Cowdrey, RN, Melissa Woollett, BHSc (Nurs);

*Auckland City Hospital, DCCM:* Colin McArthur, FJFICM, Thomas Hills, DPhil, Lynette Newby, MN, Yan Chen, MN, Catherine Simmonds, PGDipHSc, Rachael McConnochie, MN, Caroline O'Connor;

*Christchurch Hospital:* Jay Ritzema Carter, PhD, Seton Henderson, MD, Kymbalee Van Der Heyden, BSc, Jan Mehrtens, PGCert, Anna Morris, BN, Stacey Morgan, BN;

*Middlemore Hospital:* Tony Williams, BMedSc, Alex Kazemi, BMedSc, Susan Morpeth, PhD, Rima Song, PGDip, Vivian Lai, MHSc, Dinuraj Girijadevi, PGCert;

*North Shore Hospital:* Robert Everitt, FACEM, Robert Russell, BSc(Hons), Danielle Hacking, PGDipNurs;

*Rotorua Hospital:* Ulrike Buehner, FCARCSI, Erin Williams, MSc;

*Tauranga Hospital:* Troy Browne, FCICM, Kate Grimwade, FRACP, Jennifer Goodson, RN, Owen Keet, FANZCA, Owen Callender, FANZCA;

*Waikato Hospital:* Robert Martynoga, FCICM, Kara Trask, PGCert, Amelia Butler, PGCert,

*Wellington Hospital:* Paul Young, PhD, Chelsea Young, PGDip, Eden Lesona, MNSc, Shaanti Olatunji, MClinIm, Leanlove Navarra, BSc (Nurs), Raulle Sol Cruz, BSc (Nurs)

*Whangarei Hospital:* Katherine Perry, FJFICM, Ralph Fuchs, FANZCA, Bridget Lambert, PGDip;

*Taranaki Base Hospital:* Jonathan Albrett, FCICM, Carolyn Jackson, RN, Simon Kirkham, PGDip;

***Portugal***:

*Hospital de Abrantes:* Nuno José Teodoro Amaro dos Santos Catorze, MD, Tiago Nuno Alfaro Lima Pereira, MD, Ricardo Manuel Castro Ferreira, RN, Joana Margarida Pereira Sousa Bastos, PharmD, Teresa Margarida Oliveira Batista, RN;

***Romania***:
*"Dr. Victor Babes" Clinical Hospital of Infectious and Tropical Diseases Bucharest:* Simin Aysel Florescu, PhD, Delia Stanciu, MD, Mihaela Florentina Zaharia, MD, Alma Gabriela Kosa, MD, Daniel Codreanu;

***Saudi Arabia***:
*King Abdulaziz Medical City- Riyadh:* Yaseen Marabi, ATSF, Eman Al Qasim, MSN, Mohamned Moneer Hagazy, MD, Lolowa Al Swaidan, MSc, Hatim Arishi, RN

***Spain***:
*Hospital del Mar:* Rosana Muñoz-Bermúdez, MD, Judith Marin-Corral, PhD, Anna Salazar Degracia, PhD, Francisco Parrilla Gómez, MD, Maria Isabel Mateo López;
*Reina Sofia University Hospital:* Rafael León López, MD, Jorge Rodriguez, PhD, Sheila Cárcel, Rosario Carmona, MD, Carmen de la Fuente, MD, Marina Rodriguez, MD;

***United Kingdom***:
*Aberdeen Royal Infirmary:* Callum Kaye, MBChB, Angela Allan, PGDip;
*Addenbrooke's Hospital:* Charlotte Summers, PhD, Petra Polgarova MSc;
*Alder Hey Children's NHS Foundation Trust:* Stephen J McWilliam, PhD, Daniel B Hawcutt, MD, Laura Rad, BSc(Hons), Laura O'Malley, BSc(Hons), Jennifer Whitbread, BSc(Hons);
*Alexandra Hospital Redditch:* Olivia Kelsall, MBChB, Nicholas Cowley MD, Laura Wild, BSc(Hons), Jessica Thrush, RGN, Hannah Wood, BSc(Hons), Karen Austin, RGN;
*Altnagelvin Hospital:* Adrian Donnelly, FFICM, Martin Kelly, MD, Naoise Smyth MB ChB, Sinéad O'Kane, BSc(Hons), Declan McClintock, MSc, Majella Warnock, MPharm, Ryan Campbell BSc, Edmund McCallion MPharm;
*Antrim Area Hospital:* Paul Johnson, FFARCSI, Shirley McKenna, MSc, Joanne Hanley, MSc, Andrew Currie, MSc, Barbara Allen, MPharm, Clare Mc Goldrick, MPhil, Moyra Mc Master, RGN, ;
*Barnet Hospital:* Rajeev Jha, MD, Michael Kalogirou, MD, Christine Ellis, PhD, Vinodh Krishnamurthy, PhD, Vashish Deelchand, MSc, Aibhilin O'Connor, MSc;
*Basildon Universty Hospital: Dipak Mukherjee, MD, Agilan Kaliappan, MD, Anirudda Pai, MD,Mark Vertue, Anne Nicholson, Joanne Riches, Gracie Maloney, Lauren Kittridge, Amanda Solesbury, Kezia Allen*
*Belfast Health and Social Care Trust (Belfast City Hospital, Mater Infirmorium, Royal Victoria Hospital):* Jon Silversides, PhD, Peter McGuigan, MBBCh, Kathryn Ward, BSc, Aisling O'Neill, BSc, Stephanie Finn, BSc;
*Brighton and Sussex University Hospitals Trust:* Barbara Phillips, Laura Oritz-Ruiz de Gordoa, BSc;
*Bristol Royal Infirmary:* Jeremy Bewley, MBChB, Matthew Thomas, MBChB, Katie Sweet, BSc(Hons), Lisa Grimmer, BSc(Hons), Rebekah Johnson, BSc(Hons);
*Calderdale and Huddersfield Foundation Trust:* Jez Pinnell, MD, Matt Robinson, BSc(Hons), Lisa Gledhill, MSc, Tracy Wood, BSc(Hons);
*Cardiff and Vale University Health Board:* Matt Morgan, PhD, Jade Cole, BSc, Helen Hill, BSc, Michelle Davies, BN, Angharad Williams, BSc, Emma Thomas, BSc, Rhys Davies, BSc, Matt Wise, DPhil;
*Charing Cross Hospital:* David Antcliffe, PhD, Maie Templeton, MSc, Roceld Rojo, BSN, Phoebe Coghlan, MA, Joanna Smee, BSc;
*Chesterfield Royal Hospital:* Euan Mackay, MD, Jon Cort, MD, Amanda Whileman, BSc, Thomas Spencer, Nick Spittle, Sarah Beavis, MD, Anand Padmakumar, MD, Katie Dale, BSc, Joanne

Hawes, BSc, Emma Moakes, BSc, Rachel Gascoyne, BSc, Kelly Pritchard, BSc, Lesley Stevenson, BSc, Justin Cooke, MD, Karolina Nemeth-Roszpopa, MD;

*The Christie NHS Foundation Trust:* Vidya Kasipandian, FFICM, Amit Patel, Suzanne Allibone, Roman Mary-Genetu, BSc;

*Colchester Hospital:* Mohamed Ramali, FRCA, Ooi HC, MRCEM, Alison Ghosh, RN, Rawlings Osagie, PharmD, Malka Jayasinghe Arachchige, MBBS, Melissa Hartley, MBBS;

*Countess of Chester Hospital:* Peter Bamford, FFICM, Emily London, MBChB, Kathryn Cawley, MRes, Maria Faulkner, BSc, Helen Jeffrey, DipNS;

*Croydon University Hospital:* Ashok Sundar Raj, MD, Georgios Tsinaslanidis, MD, Reena Nair Khade, BSc, Gloria Nwajei Agha, BSc, Rose Nalumansi Sekiwala;

*Cumberland Infirmary:* Tim Smith, FRCA, Chris Brewer, BPharm(Hons), Jane Gregory, BSc(Hons);

*Darlington Memorial Hospital:* James Limb, FRCA, Amanda Cowton, BSc(Hons), Julie O'Brien, DipNurs, Kelly Postlethwaite, DipNurs;

*Derriford Hospital:* Nikitas Nikitas, PhD, Colin Wells, MSc, Liana Lankester, PGCert, Helen McMillan, MSc;

*Dorset County Hospital:* Mark Pulletz, FFICM, Patricia Williams, AdDip, Jenny Birch, BA, Sophie Wiseman, Mpharm, Sarah Horton, BA(Hons);

*East Kent Hospitals (Queen Elizabeth the Queen Mother Hospital):* Ana Alegria, CCT, Salah Turki, MBBch, Tarek Elsefi, MRCP, Nikki Crisp, BSc, Louise Allen, BSc;

*East Lancashire Hospitals NHS Trust (Royal Blackburn Hospital):* Matthew Smith, MD, Sri Chukkambotla, MD, Wendy Goddard, BSc, Stephen Duberley BSc;

*Freeman Hospital and Royal Victoria Infirmary, Newcastle upon Tyne*: Iain J McCullagh, FRCA, Philip Robinson, MSc, Bijal Patel, MSc, Sinead Kelly, PGDip;

*Frimley Health NHS Foundation Trust:* Omar Touma, MD, Susan Holland, Christopher Hodge, Holly Taylor, Meera Alderman, Nicky Barnes, Joana Da Rocha, BSc, Catherine Smith, BSc, Nicole Brooks, Thanuja Weerasinghe, BSc, Julie-Ann Sinclair, Yousuf Abusamra, MD, Ronan Doherty, MD, Joanna Cudlipp, MD, Rajeev Singh, MD, Haili Yu, MD, Admad Daebis, MD, Christopher NG, MD, Sara Kendrick, MD, Anita Saran, MD, Ahmed Makky, MD, Danni Greener, MD, Louise Rowe-Leete, Dip, Alexandra Edwards, Dip, Yvonne Bland, BSc, Rozzie Dolman, BSc, Tracy Foster, BSc;

*Gateshead Health NHS Trust:* Vanessa Linnett, MD, Amanda Sanderson, Jenny Ritzema, Helen Wild;

*George Eliot Hospital:* Divya Khare, FRCA, Meredith Pinder, BSN, Selvin Selvamoni, MSc, Amitha Gopinath, MBA;

*Glan Clwyd Hospital:* Richard Pugh, FFICM, Daniel Menzies, FRCP, Richard Lean, MBChB, Xinyi Qiu, MBChB, Jeremy James Scanlon, MBChB;

*Glasgow Royal Infirmary:* Kathryn Puxty, MD, Susanne Cathcart, BSc, Chris Mc Govern, MBChB, Samantha Carmichael, MRPharms, Dominic Rimmer, BSc;

*Glenfield Hospital Leicester:* Hakeem Yusuff, FFICM, Graziella Isgro, FFICM, Chris Brightling, PhD, Michelle Bourne, BSc(Hons), Michelle Craner, DipHE, Rebecca Boyles, BSc (Hons);

*Great Western Hospitals NHS Foundation Trust:* Malcolm Watters, MBBCh, Rachel Prout, MBChB, Louisa Davies, BSc, Suzannah Pegler, BSc(Hons), Lynsey Kyeremeh, BPharm, Aiman Mian, MBBS;

*Guy's & St Thomas' NHS Foundation Trust:* Manu Shankar-Hari, PhD, Marlies Ostermann, PhD, Marina Marotti, BSc, Neus Grau Novellas, BSc, Aneta Bociek, BSc;

*Hammersmith Hospital:* Stephen Brett, MD, Sonia Sousa Arias, BSc, Rebecca Elin Hall, BN;

*Homerton University Hospital NHS Foundation Trust:* Susan Jain, MD, Abhinav Gupta MD, Catherine Holbrook

*James Cook University Hospital:* Jeremy Henning, MB, Stephen Bonner, BSc, Keith Hugill, BSc, Emanuel Cirstea, MSc, Dean Wilkinson, BSc, Jessica Jones, BSc;

*James Paget University Hospitals:* Michal Karlikowski, MD, Helen Sutherland, BSc(Hons), Elva Wilhelmsen, DipHE, Jane Woods, BSc, Julie North, BSc(Hons);

*Kettering General Hospital:* Dhinesh Sundaran, FFICM, Laszlo Hollos, FFICM, Susan Coburn, PGCert, Anna Williams, BSc, Samantha Saunders, BTEC;

*King's College Hospital (Denmark Hill site):* Phil Hopkins, MD, John Smith, RN, Harriet Noble, RN, Maria Theresa Depante, RN, Emma Clarey, RN;

*Lancashire Teaching Hospitals NHS Foundation Trust:* Shondipon Laha, FFICM, Mark Verlander, MBA, Alexandra Williams, MSc;

*Leeds Teaching Hospitals Trust:* Elankumaran Paramasivam, FRCP, Elizabeth Wilby, BSc (Hons), Bethan Ogg, BSc (Hons), Clare Howcroft, BSc (Hons), Angelique Aspinwall, BSc (Hons), Sam Charlton, BSc (Hons), Richard Gould, MBBS, Deena Mistry, MPharm, Sidra Awan, MPharm, Caroline Bedford, MPharm;

*Leicester General Hospital:* Andrew Hall, MRCP, Jill Cooke, RGN, Caroline Gardiner-Hill, RGN, Carolyn Maloney, Nigel Brunskill, PhD;

*Leicester Royal Infirmary:* Hafiz R Qureshi, MRCPI, Neil Flint, MBChB, Sarah Nicholson, Sara Southin, Andrew Nicholson, Amardeep Ghattaoraya, MSc;

*Lewisham and Greenwich NHS Trust:* Dr Daniel Harding, MD, Sinead O'Halloran , Amy Collins Emma Smith , Estefania Trues;

*Liverpool Foundation Trust Aintree:* Barbara Borgatta, PhD, Ian Turner-Bone, DipHE, Amie Reddy, Laura Wilding, DipHE;

*Liverpool Heart and Chest Hospital:* Craig Wilson, Zuhra Surti;

*Luton and Dunstable University Hospital:* Loku Chamara Warnapura, FFICM, Ronan Agno, BSc, Prasannakumari Sathianathan, MSc, Deborah Shaw, FFICM, Nazia Ijaz, FFICM, Dean Burns, MD, Mohammed Nisar, MD, Vanessa Quick, MD, Craig Alexander, BSc, Sanil Patel BSc, Nafisa Hussain, Yvynne Croucher, BSc, Eva-Maria Lang, MD, Banu Rudran, MD, Syed Gilani, MD, Talia Wieder, MD, Margaret Louise Tate, BSc;

*Maidstone and Tunbridge Wells NHS Trust:* David Golden, FFICM, Miriam Davey, PGDip, Rebecca Seaman BSc (Hons);

*Manchester Royal Infirmary:* Tim Felton, FFICM, Jonathan Bannard-Smith, FFICM, Joanne Henry, Richard Clark, DiPHE, Kathrine Birchall, BSc(Hons), Joanne Henry, MA, Fiona Pomeroy, BSc (Hons), Rachael Quayle, Dip.HE, Katharine Wylie, MSc, Anila Sukuraman, BSc, John McNamarra, MD;

*Medway Maritime Hospital:* Arystarch Makowski, PhD, Beata Misztal, PhD, Iram Ahmed, PhD, Kevin Neicker, MBA, Sam Millington, BMBS, Rebecca Squires, BSc, Masroor Phulpoto, MBBS;

*Milton Keynes University Hospital:* Richard Stewart, Esther Mwaura, BSc, Louise E Mew, BSc(Hons), Lynn Wren, BSc(Hons), Felicity Willams, PhD;

*Mid & South Essex NHS Foundation Trust:* Aneta Oborska, FRCA, Rino Maeda, MBBS, Selver Kalchko-Veyssal, MD, Raji Orat Prabakaran, BSc, Bernard Hadebe, MSc, Eric Makmur, MBBS, Guy Nicholls, MBBS;

*Musgrove Park Hospital:* Richard Innes, MBBCh, Patricia Doble, BSc(Hons), Libby Graham, RN, Charmaine Shovelton, RN;

*Nevill Hall Hospital:* Vincent Hamlyn, MBBCh, Nancy Hawkins, PhD, Anna Roynon-Reed, MSc, Sean Cutler, MSc, Sarah Lewis, MBChB;

*Newham University Hospital:* Juan Martin Lazaro, PhD, Tabitha Newman, MSc;

*Ninewells Hospital:* Pauline Austin, MBChB, Susan Chapman, MBChB, Louise Cabrelli, BSc;

*Norfolk and Norwich University Hospital:* Simon Fletcher, FFICM, Jurgens Nortje, FFICM, Deirdre Fottrell-Gould, Dip, Georgina Randell, Dip, Katie Stammers, BSc;
*Northampton General Hospital:* Mohsin Zaman, MRCP, Einas Elmahi, MPhil, Andrea Jones, PhD, Kathryn Hall, Dip;
*Northern General Hospital, Sheffield:* Gary H Mills, PhD, Kim Ryalls, RegPharmTech, Kate Harrington RCN, Helen Bowler, BSc, Jas Sall, BSc, Richard Bourne, PhD;
*North Manchester General Hospital:* Zoe Borrill, MD, Tracy Duncan, MD, Thomas Lamb, MD, Joanne Shaw, BSc, Claire Fox, BSc, Kirstie Smith, BSc, Sarah Holland, Bethany Blackledge, BSc, Liam McMorrow, BSc, Laura Durrans, Jade Harris;
*North Middlesex University Hospital:* Jeronimo Moreno Cuesta, MD, Kugan Xavier, MD, Dharam Purohit, EDIC, Munzir Elhassan, MBBS, Anne Haldeos, BSc, Rachel Vincent, DipHE, Marwa Abdelrazik, MBBCH, Samuel Jenkins, BMBS, Arunkumar Ganesan, MD, Rohit Kumar, DA, David Carter, MBBS, Dhanalakshmi Bakthavatsalam, BSc;
*Oxford University Hospitals:* Matthew Rowland, FFICM, Paula Hutton, PGCert, Archana Bashyal, MSc, Neil Davidson, BSc, Clare Hird, MSc, Sally Beer, MSc;
*Pilgrim Hospital Boston:* Manish Chhablani, FFICM, Gunjan Phalod, MPharm, Amy Kirkby, BSc(Hons), Simon Archer, BSc(Hons), Kimberley Netherton, RGN;
*Princess Royal Hospital:* Barbara Philips, MD, Dee Mullan, BSc, Denise Skinner, BSc, Jane Gaylard, BSc, Julie Newman, BSc;
*Princess of Wales Hospital:* Sonia Arun Sathe, MD, Lisa Roche, BSc, Ellie Davies, BSc, Keri Turner;
*Poole Hospital:* Henrik Reschreiter, FFICM, Julie Camsooksai, PGDE, Sarah Patch, BSc(Hons), Sarah Jenkins, BSc(Hons), Charlotte Humphrey, BSc (Hons);
*Queen Alexandra Hospital Portsmouth:* David Pogson, MSc, Steve Rose, BSc, Zoe Daly, BSc, Lutece Brimfield, BN, Angie Nown;
*Queen Elizabeth Hospital, Birmingham:* Dhruv Parekh, PhD, Colin Bergin, BSc, Michelle Bates, BSc, Christopher McGhee, BSc, Daniella Lynch, BSc, Khushpreet Bhandal, Dip, Kyriaki Tsakiridou, MSc, Amy Bamford, BSc, Lauren Cooper, MSc, Tony Whitehouse, MD, Tonny Veenith, MD;
*Queen Elizabeth University Hospital, Glasgow:* Malcolm A.B. Sim, MD, Sophie Kennedy Hay, BN, Steven Henderson, MPH, Maria Nygren, MSc, Eliza Valentine, HNC;
*Queen's Hospital, Burton:* Amro Katary, MD, Gill Bell, BSc, Louise Wilcox, BSc, Katy English BSc, Ann Adams;
*Queen's Hospital, Romford:* Mandeep-Kaur Phull, MBBS, Abbas Zaidi, MBBS, Tatiana Pogreban, BN, Lace Paulyn Rosaroso, BN;
*Queens Medical Centre and Nottingham City Hospital:* Daniel Harvey, BMBS, Benjamin Lowe, BMBS, Megan Meredith, BSc(Hons), Lucy Ryan, MNSc, DREEAM Research Team;
*The Rotherham NHS Foundation Trust:* Anil Hormis, FRCA, Rachel Walker, BA, Dawn Collier, BSc, Sarah Kimpton, MSc, Susan Oakley;
*Royal Alexandra Hospital:* Kevin Rooney, MBChB, Natalie Rodden, BSc, Emma Hughes, BSc, Nicola Thomson, BSc(Hons), Deborah McGlynn, BSc, Charlotte Clark, Dip, Patricia Clark, BSc;
*Royal Berkshire Hospital:* Andrew Walden, FFICM, Liza Keating, MBChB, Matthew Frise, DPhil, Tolu Okeke, BSc, Nicola Jacques, MSc, Holly Coles, BSc, Emma Tilney, BSc, Emma Vowell, DipHE;
*Royal Bournemouth and Christchurch Hospitals:* Martin Schuster-Bruce, FRCA, Sally Pitts, BSc, Rebecca Miln, ADipHE, Laura Purandare, MBA, Luke Vamplew, BSc;
*Royal Brompton Hospital:* Brijesh Patel, FRCA, Debra Dempster, Mahitha Gummadi, Natalie Dormand, Shu Fang Wang;
*Royal Cornwall NHS Trust:* Michael Spivey, FFICM, Sarah Bean, RN, Karen Burt, RN, Lorraine Moore, MPharm;

*Royal Devon and Exeter NHS Foundation Trust:* Christopher Day, MD, Charly Gibson, MBChB, Elizabeth Gordon, BSc, Letizia Zitter, BSc, Samantha Keenan, BSc;

*Royal Glamorgan Hospital:* Jayaprakash Singh, MD, Ceri Lynch, MD, Lisa Roche, Justyna Mikusek, Bethan Deacon, Keri Turner;

*Royal Gwent Hospital*: Tamas Szakmany, PhD, Evelyn Baker, MSc, Shiney Cherian, BSc(Hons), John Hickey, MSc, Shreekant Champanerkar, MBBS ;

*Royal Hallamshire Hospital, Sheffield:* Gary H Mills, PhD, Ajay Raithatha, FFICM, Kris Bauchmuller, FFICM, Norfaizan Ahmad, FFICM, Matt Wiles FFICM, Jayne Willson, RN;

*Royal Hampshire Hospitals:* Irina Grecu, MD, Jane Martin, Caroline Wrey Brown, Ana-Marie Arias, Emily Bevan;

*Royal Infirmary of Edinburgh:* Thomas H Craven, PhD, David Hope, PGDip, Jo Singleton, BN, Sarah Clark, MNurs, Corrienne McCulloch, PhD;

*Royal Liverpool University Hospital:* Ingeborg D Welters, PhD, David Oliver Hamilton, BMBS, Karen Williams, RGN, Victoria Waugh, BA, David Shaw, DipHE, Suleman Mulla, MBChB, Alicia Waite, PhD, Jaime Fernandez Roman, BSc, Maria Lopez Martinez, BSc;

*Royal London Hospital:* Zudin Puthucheary, PhD, Timothy Martin, BA(Hons), Filipa Santos, RN, Ruzena Uddin, MSc(Hons), Maria Fernandez, MSc, Fatima Seidu, MSc, Alastair Somerville, MSc, Mari Lis Pakats, MSc, Priya Dias, PhD, Salam Begum, BSc, Tasnin Shahid, BSc;

*The Royal Free Hospital:* Sanjay Bhagani, FRCP, Mark De Neef, MSc, Helder Filipe, BSc, Sara Mingos, BSc, Amitaa Maharajh, BA, Glykeria Pakou, BA, Aarti Nandani, MPharm;

*The Royal Marsden NHS Foundation Trust:* Kate Colette Tatham, PhD, Shaman Jhanji, PhD, Ethel Black, BSNurs, Arnold Dela Rosa, BSNurs, Ryan Howle, FRCA, Ravishankar Rao Baikady, FRCA;

*The Royal Oldham Hospital:* Redmond P Tully, FFICM, Andrew Drummond, FFICM, Joy Dearden, BSc, Jennifer E Philbin, MSc, Sheila Munt, SRN;

*The Royal Wolverhampton NHS Trust:* Shameer Gopal, MBBCh, Jagtar- Singh Pooni, MBBS, Saibal Ganguly, MBBS, Andrew Smallwood, RGN, Stella Metherell, RGN;

*Royal Papworth Hospital:* Alain Vuylsteke, MD, Charles Chan, FRCA, Saji Victor, COVID Research Team, Papworth Hospital;

*Royal Stoke Hospital:* Ramprasad Matsa, FRCP, Minerva Gellamucho, BSN, Michelle Davies, NVQ;

*Royal Surrey County Hospital:* Ben Creagh-Brown, PhD, Joe Tooley, MSc, Laura Montague, BSc, Fiona De Beaux, BSc, Laetitia Bullman, MBChB;

*Royal United Hospital Bath:* Ian Kerslake, FFICM, Carrie Demetriou, RN, Sarah Mitchard, MBBS, Lidia Ramos, RN, Katie White, MSc;

*Russells Hall Hospital:* Michael Reay, FFICM, Steve Jenkins, MD, Caroline Tuckwell, Angela Watts, BSc, Eleanor Traverse, Stacey Jennings;

*Salisbury NHS Foundation Trust:* Phil Donnison, FFICM, Maggie Johns, RGN, Ruth Casey, BSc, Lehentha Mattocks, Dip, Sarah Salisbury;

*Salford Royal NHS Foundation Trust:* Paul Dark, PhD, Alice Harvey, BSc, Reece, Doonan, BSc, Liam McMorrow, BA (Hons), Karen Knowles, BA (Hons);*Sandwell and West Birmingham NHS Trust:* Jonathan Hulme, FFICM, Santhana Kannan, FFICM, Sibet Joseph, BSc, Fiona Kinney, RGN, Ho Jan Senya, BPharm;

*Sherwood Forest Hospitals NHS Foundation Trust:* Valli Ratnam, MD, Mandy Gill, Jill Kirk, Sarah Shelton

*South Tyneside District Hospital:* Christian Frey, MD, Riccardo Scano, MD, Madeleine McKee, BSc, Peter Murphy, BSc;

*Southmead Hospital:* Matt Thomas, FFICM, Ruth Worner, RGN, Beverley Faulkner, RGN, Emma Gendall, BSc, Kati Hayes, BSc, Hayley Blakemore, BSc, Borislava Borislavova, MSc;

*St. Bartholomew's Hospital:* Colin Hamilton-Davies, MBBS, Carmen Chan, BSc, Celina Mfuko, BSc, Hakam Abbass, MSc, Vineela Mandadapu, MSc;

*St. George's Hospital:* Susannah Leaver, MRCP, Kamal Patel, MRCP, Sarah Farnell-Ward, MSc, Romina Pepermans Saluzzio, BSc, John Rawlins, MBBS;

*St. Mary's Hospital:* Anthony Gordon, MD; Dorota Banach, BSc, Ziortza Fernández de Pinedo Artaraz, BN, Leilani Cabreros, BSN;

*St. Peter's Hospital, Chertsey:* Ian White, FFICM, Maria Croft, BSc(Hons), Nicky Holland, BN(Hons), Rita Pereira, MPharm;

*Stepping Hill Hospital, Stockport:* Ahmed Zaki, PhD, David Johnson, MPhil, Matthew Jackson, MBChB, Hywel Garrard, BMBS, Vera Juhaz, MD, Louise Brown BSc (Hons);

*Sunderland Royal Hospital:* Alistair Roy, MBChB, Anthony Rostron, PhD, Lindsey Woods, BSc, Sarah Cornell, BSc;

*Swansea Bay University Health Board:* Suresh Pillai, FFCIM, Rachel Harford, RN, Helen Ivatt, FRCA, Debra Evans, BN, Suzanne Richards, BN, Eilir Roberts, MBBCh, James Bowen, MBBCh James Ainsworth, MBBS;

*Torbay and South Devon NHS Foundation Trust:* Thomas Clark, MBChB, FRCA, FFICM, Angela Foulds, BSc, Stacey Atkins, Dip RN;

*United Lincolnshire NHS Trust:* Kelvin Lee, PhD, Russell Barber, FRCA, Anette Hilldrith, RGN, Claire Hewitt, RGN, Gunjan Phalod, MPharm;

*University Hospitals Coventry & Warwickshire NHS Trust:* Pamela Bremmer, BSc, Geraldine Ward, MA, Christopher Bassford, PhD;

*University Hospital of North Tees:* Farooq Brohi, FFARCSI, Vijay Jagannathan, FRCA, Michele Clark, MA, Sarah Purvis, Dip, Bill Wetherill, MSc;

*University Hospital Southampton NHS Foundation Trust:* Ahilanandan Dushianthan, PhD, Rebecca Cusack, MD, Kim de Courcy-Golder, PGDip, Karen Salmon, MSc, Rachel Burnish, Simon Smith, BN, Susan Jackson, BSc, Winningtom Ruiz, BSc (Hons), Zoe Duke BSc (Hons) Magaret Johns, BA, Michelle Male, Dip, Kirsty Gladas, BSc (Hons), Satwinder Virdee, MPharm, Jacqueline Swabe, MPharm, Helen Tomlinson;

*Warwick Hospital:* Ben Attwood, MBBCh, Penny Parsons, BSc, Bridget Campbell, BSc, Alex Smith, BSc;

*Watford General Hospital:* Valerie J Page, MBBCh, Xiao Bei Zhao, BSc (Hons), Deepali Oza, BPharm, Gail Abrahamson, DPhil, Ben Sheath, BSc (Hons), Chiara Ellis, BSc (Hons);

*Western General Hospital, Edinburgh:* Jonathan Rhodes, PhD, Thomas Anderson, MBChB, Sheila Morris;

*Whipps Cross Hospital:* Charlotte Xia Le Tai, MBChB, Amy Thomas, MSc, Alexandra Keen, MSc;

*Whiston Hospital*: Dr Ascanio, Tridente, Karen Shuker, Jeanette Anders, Sandra Greer, Paula Scott, Amy Millington, Philip Buchanan, Jodie Kirk

*Wirral University Teaching Hospital NHSFT:* Craig Denmade, MBChB, Girendra Sadera, MBBS, Reni Jacob, BSc, Cathy Jones, BSc, Debbie Hughes;

*Worcester Royal Hospital:* Stephen Digby, MBBS, Nicholas Cowley, MD, Laura Wild, BSc(Hons), Jessica Thrush, RGN, Hannah Wood, BSc(Hons), Karen Austin, RGN;

*Wrexham Maelor Betsi Cadwaladr University Hospital*: David Southern, FFICM, Harsha Reddy, FFICM, Sarah Hulse, BSc, Andy Campbell, FFICM, Mark Garton, Claire Watkins, PGDip, Sara Smuts, BN;

*Wrightington, Wigan and Leigh Teaching Hospitals NHS Foundation Trust:* Alison Quinn, MD, Benjamin Simpson, MD, Catherine McMillan, MD, Cheryl Finch, BSc, Claire Hill BSc, Josh Cooper;

*Wye Valley NHS Trust:* Joanna Budd, MBBS, Charlotte Small, PhD, Ryan O'Leary, MBBS, Janine Birch, RN, Emma Collins, BSc (Hons);
*Wythenshawe Hospital*: Peter D G Alexander, FFICM, Tim Felton, FFICM, Susan Ferguson, BSc, Katharine Sellers, BSc, Joanne Bradley-Potts, BSc;
*York Teaching Hospital*: David Yates, FCRA, Isobel Birkinshaw, BSc(Hons), Kay Kell, BSc(Hons), Zoe Scott, BN, Harriet Pearson, BSc;

***United States of America***
*University of Pittsburgh Medical Center*: David Huang, MD, Tim Girard, MD, Kim Basile, BS, Erin McCreary, PharmD, Kelsey Linstrum, MS;

### 2. Additional methods

#### *2.1 REMAP-CAP Eligibility Criteria for suspected or proven Covid-19*

*Platform inclusion criteria:*

- Adult patient admitted to hospital with acute illness due to suspected or proven pandemic (Covid-19) infection

*Platform exclusion criteria:*

- Death is deemed to be imminent and inevitable during the next 24 hours AND one or more of the patient, substitute decision maker or attending physician are not committed to full active treatment
- Patient is expected to be discharged from hospital today or tomorrow
- More than 14 days have elapsed while admitted to hospital with symptoms of an acute illness due to suspected or proven pandemic infection
- Previous participation in this REMAP within the last 90 days

*Immune Modulation Therapy domain specific inclusion criteria:*

- Severe disease state, defined by receiving respiratory or cardiovascular organ failure support in an intensive care unit (ICU).
    a. Respiratory organ support is defined as invasive or non-invasive mechanical ventilation including via high flow nasal cannula if flow rate >30 L/min and $F_IO_2$ >0.4. If non-invasive ventilation would normally be provided but is being withheld, due to infection control concerns associated with aerosol generating procedures, then the patient still meets the severe disease state criteria.
    b. Cardiovascular organ support was defined as the intravenous infusion of any vasopressor or inotrope.
    c. Pandemic surge capacity means that provision of advanced organ support may need to occur in locations that do not usually provide ICU-level care. Therefore, an ICU is defined as an area within the hospital that is repurposed so as to be able to deliver one or more of the qualifying organ failure

supports (non-invasive ventilation, invasive ventilation, and vasopressor therapy)

- Microbiological testing for SARS-CoV-2 of upper or lower respiratory tract secretions or both has occurred or is intended to occur

*Immune Modulation Therapy domain specific exclusion criteria:*

- More than 24 hours has elapsed since ICU admission
- Patient has already received any dose of one or more of any form of interferon, anakinra, tocilizumab, or sarilumab during this hospitalization or is on long-term therapy with any of these agents prior to this hospital admission
- Known condition or treatment resulting in ongoing immune suppression including neutropenia prior to this hospitalization
- Patient has been randomized in a trial evaluating an immune modulation agent for proven or suspected Covid-19 infection, where the protocol of that trial requires ongoing administration of study drug
- The treating clinician believes that participation in the domain would not be in the best interests of the patient

*Intervention specific exclusion criteria:*

- Known hypersensitivity to an agent specified as an intervention in this domain will exclude a patient from receiving that agent
- Intention to prescribe systemic corticosteroids for any reason, other than participation in the Corticosteroid domain of this platform, will result in exclusion from receiving IFN-β1a
- Known hypersensitivity to proteins produced by E. coli will result in exclusion from receiving anakinra
- Known or suspected pregnancy will result in exclusion from the anakinra, IFN-β1a, tocilizumab, and sarilumab interventions. It is normal clinical practice that women admitted who are in an age group in which pregnancy is possible will have a pregnancy test conducted. The results of such tests will be used to determine interpretation of this exclusion criteria.

- A baseline alanine aminotransferase or an aspartate aminotransferase that is more than five times the upper limit of normal will result in exclusion from receiving tocilizumab or sarilumab
- A baseline platelet count < 50 x $10^9$ / L will result in exclusion from receiving tocilizumab or sarilumab

#### *2.2 Additional Statistical Methods:*

##### *Secondary outcomes:*

- Hospital mortality / survival (dichotomous)
- 90-day survival (time to event)
- Respiratory support-free days
- Cardiovascular support-free days
- Time to ICU discharge
- Time to hospital discharge
- WHO ordinal scale (range: 0-8, where 0 = no illness, 1-7 = increasing level of care, and 8 = death) assessed at day 14.[1] In this analysis categories 0,1,2 have been condensed into one category for all patients discharge from hospital.
- Progression to invasive mechanical ventilation, extracorporeal membrane oxygenation (ECMO), or death among those not ventilated at baseline

##### *Analysis populations:*

The primary analysis was conducted on all severe state patients with Covid-19 randomized to any domain allowing maximal incorporation of all information, providing the most robust estimation of the coefficients of all covariates. As the primary model included information about assignment to interventions within domains whose evaluation is ongoing and blinded to the investigators, the primary analysis was run by the fully unblinded Statistical Analysis Committee, who conduct all protocol-specified trial update analyses and report those results to the DSMB.

In addition to the primary analysis conducted by the Statistical Analysis Committee, analysis of the primary outcome was then repeated in a second model using only data from those patients enrolled in those domains that had ceased recruitment, Corticosteroid domain and Covid-19 Antiviral domain, as well as those patients randomised in the Immune Modulation Therapy domain; the Unblinded Intention To Treat (ITT) population. For this analysis there was no adjustment for assignment to interventions in other ongoing domains. These analyses were conducted by the investigator team who remain blinded to on-going domains and interventions in the platform.

Further secondary analyses explored the effects of excluding patients who were ruled out for Covid-19 (defined as documented negative tests for SARS-CoV-2 infection and no positive tests), including only those patients randomised directly to tocilizumab or sarilumab versus control. Additional analyses conducted by the investigator team included analyzing only patients randomized to tocilizumab or sarilumab and corresponding controls without any interactions from other domains (IL-6 RA specific ITT population) and a subset of this population treated as per protocol (IL-6 RA per protocol population)

One subgroup was defined in the statistical analysis plan, dividing patients into terciles according to serum C-reactive protein levels at study inclusion.

Various sensitivity analyses were conducted including less informative standard normal priors on pre-specified interactions excluding adjustment for site and time epoch, and alternative coding of steroid interventions after closure of the corticosteroid domain. Identical analyses were conducted to estimate the effect on mortality, except the outcome was dichotomous (alive or dead at hospital discharge). The secondary outcomes were also analysed in the unblinded ITT model. The time-to-death and length of stay outcomes were time-to-event analyses with results expressed as hazard ratios. The primary safety analysis compared the proportion of patients who developed one or more serious adverse events across groups.

#### Additional Results

##### *Site Participation in the Immune Modulation Domain*

During the study period, April 19 to November 19, 2020, 125 sites were open for enrollment in the Immune Modulation Domain. There were 113 sites open to randomisation for tocilizumab and / or sarilumab, 98 sites in the UK, 7 in The Netherlands, 3 in Australia, 2 in New Zealand, 2 in Ireland and 1 in the Kingdom of Saudi Arabia. The trial was approved by the following independent ethics committees in these countries; UK - London-Surrey Borders Research Ethics Committee; Netherlands - Medisch Ethische Toetsingscommissie Utrecht (METC Utrecht); Australia - Sydney Local District ethics Review Committee (Royal Prince Alfred Hospital); New Zealand – Northern A Health and Disability Ethics Committee; Ireland – St Vincent's Healthcare Group Ethics and Medical Research Committee; Kingdom of Saudi Arabia – King Abdullah International Medical Research Center Institutional Review Board.

##### *Platform exclusions:*

From the patient flow figure 1

764 patients were excluded from the REMAP-CAP platform, severe state:

2 - Prior enrollment in REMAP-CAP within 90 days
5 - No lower respiratory signs or symptoms *
11 - No radiological infiltrate *
318 - No qualifying organ support in ICU
375 - Exceeded platform eligibility time windows
32 - Death deemed imminent and inevitable
136 - Other

* = exclusion criteria from the original core protocol for community acquired pneumonia, but dropped once Covid-19 specific amendments described in the Pandemic Appendix to

Core protocol were implemented (see protocol). Patients could be excluded for more than one reason.

##### *Additional analyses:*

The additional analyses specified in the statistical analysis plan are provided in the "REMAP-CAP Unblinded Analysis Report" supplement, with the exception of the anti-viral interactions with immune modulation interventions. The Covid-19 antiviral primary analysis has not yet been completed or reported, and these treatment interactions will be included in that subsequent report. Provided below is the sensitivity analysis of the primary outcome (organ support-free days and hospital survival, with no treatment interactions included and the pre-specified C-reactive protein subgroup analysis (Table S1). The cut points of CRP terciles were 102 and 187 µg/ml.  There was one additional post-hoc analysis undertaken and presented that does not use any borrowing between to the two IL-6 receptor antagonists, tocilizumab and sarilumab (Table S2). In this model they are no longer "nested". Table S3 and S4 provide the odds ratios and 95% CrI for each parameter in the second model using data from those patients enrolled in those domains that had ceased recruitment, Corticosteroid domain and Covid-19 Antiviral domain, as well as those patients randomised in the Immune Modulation Therapy domain; the Unblinded ITT population.

Table S1. Additional analyses of the Primary Outcomes

| Outcome/Analysis | Tocilizumab (N=353) | Sarilumab (N=48) | Control (N=402) |
|---|---|---|---|
| **Secondary Analysis of Primary Outcome**, model restricted to the IL-6 RA population only with no treatment interactions included | | | |
| Adjusted OR - mean (SD) | 1.69 (0.23) | 1.91 (0.47) | 1 |
| - median (95% CrI) | 1.67 (1.29 to 2.18) | 1.83 (1.23 to 3.04) | 1 |
| Probability of superiority to control, % | >99.9 | 99.9 | - |
| **Secondary analysis of Hospital Survival,** model restricted to the IL-6 RA population only with no treatment interactions included | | | |
| Adjusted OR - mean (SD) | 1.72 (0.31) | 2.31 (1.01) | 1 |
| - median (95% CrI) | 1.69 (1.19 to 2.42) | 2.05 (1.22 to 5.08) | 1 |
| Probability of superiority to control, % | 99.8 | 99.7 | - |
| **Secondary Analysis of Primary Outcome**, model restricted to Immune Modulation Therapy Domain participants, according to **CRP tercile subgroups** | | | |
| **CRP lowest tercile** | | | |
| Adjusted OR - mean (SD) | 1.51 (0.42) | 1.86 (1.06) | 1 |
| - median (95% CrI) | 1.45 (0.85 to 2.48) | 1.62 (0.76 to 4.57) | 1 |
| Probability of superiority to control, % | 91.3 | 90.3 | - |
| **CRP middle tercile** | | | |
| Adjusted OR - mean (SD) | 1.54 (0.41) | 1.51 (0.54) | 1 |
| - median (95% CrI) | 1.49 (0.89 to 2.49) | 1.44 (0.73 to 2.75) | 1 |
| Probability of superiority to control, % | 93.5 | 86.6 | - |
| **CRP highest tercile** | | | |
| Adjusted OR - mean (SD) | 2.00 (0.58) | 2.60 (1.44) | 1 |
| - median (95% CrI) | 1.92 (1.12 to 3.34) | 2.25 (1.08 to 6.46) | 1 |
| Probability of superiority to control, % | 99.1 | 98.6 | - |
| **Secondary Analysis of Hospital Survival**, model restricted to Immune Modulation Therapy Domain participants, according to **CRP tercile subgroups** | | | |
| **CRP lowest tercile** | | | |
| Adjusted OR - mean (SD) | 1.60 (0.61) | 2.05 (1.86) | 1 |
| - median (95% CrI) | 1.49 (0.74 to 3.08) | 1.66 (0.71 to 5.79) | 1 |
| Probability of superiority to control, % | 86.8 | 87.5 | - |
| **CRP middle tercile** | | | |
| Adjusted OR - mean (SD) | 1.72 (0.68) | 1.82 (1.06) | 1 |
| - median (95% CrI) | 1.60 (0.77 to 3.40) | 1.61 (0.66 to 4.26) | 1 |
| Probability of superiority to control, % | 89.3 | 85.8 | - |
| **CRP highest tercile** | | | |
| Adjusted OR - mean (SD) | 1.62 (0.61) | 2.43 (3.28) | 1 |
| - median (95% CrI) | 1.51 (0.76 to 3.12) | 1.80 (0.73 to 7.88) | 1 |
| Probability of superiority to control, % | 87.4 | 90.0 | - |

Table S2. Exploratory post-hoc analyses of the Primary Outcomes

| **Outcome/Analysis** | **Tocilizumab (N=353)** | **Sarilumab (N=48)** | **Control (N=402)** |
|---|---|---|---|
| **Exploratory post-hoc Analysis of Primary Outcome**, model restricted to Immune Modulation Therapy Domain participants, with no borrowing between IL-6 RA interventions | | | |
| Adjusted OR - mean (SD) | 1.64 (0.23) | 1.98 (0.60) | 1 |
| - median (95% CrI) | 1.62 (1.24 to 2.14) | 1.90 (1.08 to 3.41) | 1 |
| Probability of superiority to control, % | >99.9 | 98.7 | - |
| **Exploratory post-hoc Analysis of Hospital Survival**, model restricted to Immune Modulation Therapy Domain participants, with no borrowing between IL-6 RA interventions | | | |
| Adjusted OR - mean (SD) | 1.58 (0.30) | 2.73 (1.13) | 1 |
| - median (95% CrI) | 1.56 (1.08 to 2.24) | 2.54 (1.17 to 5.56) | 1 |
| Probability of superiority to control, % | 99.1 | 99.2 | - |

Table S3 Odds ratios for secondary analysis of organ support free days for the unblinded ITT population

| Parameter | Mean | SD | Median | CrI |
|---|---|---|---|---|
| Age<39 | 4.11 | 0.91 | 4.01 | (2.61, 6.17) |
| Age 40-49 | 2.11 | 0.36 | 2.08 | (1.49, 2.88) |
| Age 50-59 | 1.96 | 0.27 | 1.94 | (1.48, 2.56) |
| Age 70-79 | 0.52 | 0.08 | 0.51 | (0.38, 0.69) |
| Age 80+ | 0.34 | 0.09 | 0.32 | (0.19, 0.55) |
| Female | 1.17 | 0.13 | 1.16 | (0.93, 1.44) |
| Time epoch 1 | 0.95 | 0.08 | 0.95 | (0.79, 1.11) |
| Time epoch 2 | 0.91 | 0.13 | 0.90 | (0.66, 1.17) |
| Time epoch 3 | 0.96 | 0.17 | 0.95 | (0.67, 1.33) |
| Time epoch 4 | 1.06 | 0.22 | 1.04 | (0.70, 1.56) |
| Time epoch 5 | 1.16 | 0.27 | 1.13 | (0.74, 1.78) |
| Time epoch 6 | 1.22 | 0.29 | 1.18 | (0.77, 1.88) |
| Time epoch 7 | 1.26 | 0.29 | 1.22 | (0.80, 1.92) |
| Time epoch 8 | 1.29 | 0.30 | 1.25 | (0.83, 1.98) |
| Time epoch 9 | 1.23 | 0.27 | 1.20 | (0.80, 1.88) |
| Time epoch 10 | 1.13 | 0.24 | 1.10 | (0.74, 1.69) |
| Time epoch 11 | 0.97 | 0.20 | 0.95 | (0.64, 1.41) |
| Time epoch 12 | 0.86 | 0.19 | 0.84 | (0.55, 1.27) |
| Time epoch 13 | 0.85 | 0.19 | 0.83 | (0.54, 1.26) |
| Time epoch 14 | 0.94 | 0.22 | 0.92 | (0.59, 1.43) |
| Time epoch 15 | 1.11 | 0.32 | 1.06 | (0.61, 1.86) |
| Time epoch 16 | 1.41 | 0.66 | 1.26 | (0.59, 3.08) |
| Tocilizumab | 1.68 | 0.24 | 1.66 | (1.26, 2.18) |
| Sarilumab | 1.84 | 0.44 | 1.77 | (1.18, 2.90) |
| Fixed-dose corticosteroid | 1.45 | 0.33 | 1.42 | (0.92, 2.18) |
| Shock-dependent corticosteroid | 1.15 | 0.26 | 1.12 | (0.72, 1.75) |
| Tocilizumab*Fixed-dose interaction effect | 1.00 | 0.05 | 1.00 | (0.91, 1.10) |
| Tocilizumab*Shock-dependent interaction effect | 1.00 | 0.05 | 1.00 | (0.91, 1.10) |
| Sarilumab*Fixed-dose interaction effect | 1.01 | 0.05 | 1.00 | (0.91, 1.11) |
| Sarilumab*Shock-dependent interaction effect | 1.00 | 0.05 | 1.00 | (0.91, 1.10) |
| Tocilizumab*Fixed-dose combination | 2.42 | 0.65 | 2.34 | (1.39, 3.91) |
| Tocilizumab*Shock-dependent combination | 1.93 | 0.54 | 1.86 | (1.08, 3.17) |
| Sarilumab*Fixed-dose combination | 2.68 | 0.90 | 2.53 | (1.37, 4.86) |
| Sarilumab*Shock-dependent combination | 2.11 | 0.72 | 1.99 | (1.08, 3.87) |

Table S4 Odds ratios for secondary analysis of hospital survival for the unblinded ITT population

| Parameter | Mean | SD | Median | CrI |
|---|---|---|---|---|
| Age<39 | 12.85 | 6.45 | 11.38 | (5.00, 28.81) |
| Age 40-49 | 3.33 | 0.89 | 3.21 | (1.95, 5.45) |
| Age 50-59 | 2.99 | 0.60 | 2.93 | (2.00, 4.34) |
| Age 70-79 | 0.41 | 0.07 | 0.41 | (0.29, 0.58) |
| Age 80+ | 0.23 | 0.07 | 0.22 | (0.12, 0.39) |
| Female | 1.19 | 0.19 | 1.18 | (0.87, 1.59) |
| Time epoch 1 | 0.94 | 0.08 | 0.94 | (0.78, 1.10) |
| Time epoch 2 | 0.87 | 0.14 | 0.87 | (0.61, 1.14) |
| Time epoch 3 | 0.86 | 0.17 | 0.85 | (0.55, 1.22) |
| Time epoch 4 | 0.87 | 0.20 | 0.86 | (0.53, 1.33) |
| Time epoch 5 | 0.90 | 0.23 | 0.87 | (0.53, 1.41) |
| Time epoch 6 | 0.91 | 0.24 | 0.88 | (0.53, 1.48) |
| Time epoch 7 | 0.92 | 0.25 | 0.89 | (0.52, 1.48) |
| Time epoch 8 | 0.95 | 0.25 | 0.92 | (0.55, 1.53) |
| Time epoch 9 | 0.99 | 0.26 | 0.95 | (0.58, 1.59) |
| Time epoch 10 | 1.02 | 0.27 | 0.98 | (0.60, 1.66) |
| Time epoch 11 | 1.02 | 0.27 | 0.97 | (0.60, 1.65) |
| Time epoch 12 | 0.99 | 0.26 | 0.96 | (0.58, 1.58) |
| Time epoch 13 | 1.00 | 0.28 | 0.96 | (0.57, 1.63) |
| Time epoch 14 | 1.08 | 0.33 | 1.03 | (0.57, 1.86) |
| Time epoch 15 | 1.23 | 0.50 | 1.14 | (0.55, 2.43) |
| Time epoch 16 | 1.52 | 1.03 | 1.27 | (0.50, 3.95) |
| Tocilizumab | 1.67 | 0.31 | 1.65 | (1.15, 2.34) |
| Sarilumab | 2.24 | 0.94 | 2.00 | (1.17, 4.69) |
| Fixed-dose corticosteroid | 0.99 | 0.29 | 0.95 | (0.54, 1.68) |
| Shock-dependent corticosteroid | 1.21 | 0.38 | 1.15 | (0.63, 2.10) |
| Tocilizumab*Fixed-dose interaction | 0.99 | 0.05 | 0.99 | (0.89, 1.09) |
| Tocilizumab*Shock-dependent interaction | 1.00 | 0.05 | 1.00 | (0.91, 1.11) |
| Sarilumab*Fixed-dose interaction | 1.01 | 0.05 | 1.00 | (0.91, 1.11) |
| Sarilumab*Shock-dependent interaction | 1.00 | 0.05 | 1.00 | (0.91, 1.10) |
| Tocilizumab*Fixed-dose combination | 1.63 | 0.58 | 1.53 | (0.77, 3.03) |
| Tocilizumab*Shock-dependent combination | 2.03 | 0.76 | 1.90 | (0.92, 3.87) |
| Sarilumab*Fixed-dose combination | 2.21 | 1.14 | 1.94 | (0.86, 5.21) |
| Sarilumab*Shock-dependent combination | 2.69 | 1.42 | 2.35 | (1.02, 6.35) |
